# Clonal Hematopoiesis Dynamics and Evolutionary Fitness During Cancer Treatment Impact Clinical Outcomes

**DOI:** 10.1101/2025.08.27.25334581

**Authors:** Mona Arabzadeh, Yi-Han Tang, Christelle Colin-Leitzinger, Sadegh Marzban, Daniel Walgenbach, Stefania Morganti, Vaidhyanathan Mahaganapathy, Erika Harper, Mingxiang Teng, Jacob K. Kresovich, Iman Washington, Heather A. Parsons, Judy E. Garber, Jeffrey West, Shridar Ganesan, Hossein Khiabanian, Nancy Gillis

## Abstract

Clonal hematopoiesis (CH) is the age-related expansion of mutated hematopoietic stem cells without other hematologic abnormalities. In patients with solid tumors, CH is associated with higher mortality and may evolve to therapy-related myeloid neoplasms; however, the mechanisms by which cancer treatments promote CH dynamics remain largely unknown. Here, we analyzed 392 serial samples from a prospective cohort of breast cancer patients and showed that cytotoxic treatments (chemotherapy ± radiation) led to strong therapeutic bottlenecks, resulting in significant reductions in hematopoietic allelic populations and differential clonal selection. CH clones that were positively selected and expanded through dose-dependent therapeutic bottlenecks frequently harbored mutations in *TP53, PPM1D, SRCAP, DNMT3A*, and *YLPM1*. Patients with positively selected CH during treatment had the shortest progression-free and overall survival compared to patients with unchanging or negatively selected CH across all therapies. These findings, corroborated in independent breast cancer and pan-cancer cohorts, provide strong evidence for clinical relevance of changes in CH during cancer treatment which may help identify patients at high risk for inferior outcomes.

## INTRODUCTION

Clonal hematopoiesis (CH) is caused by the expansion of hematopoietic stem cells that carry somatic alterations in genes recurrently mutated in myeloid neoplasms. CH mutations with variant allele frequencies above 2% are defined as clonal hematopoiesis of indeterminate potential (CHIP) ^1^ and are associated with an elevated risk of overall mortality, especially from cardiovascular causes and progression of or to neoplasia^2–4^. CH mutations are more common in individuals with solid tumors compared to healthy population-based cohorts^5^ and are routinely detected in blood and tumor specimens^6–8^.

The growth pattern and Darwinian evolution of mutation-driven CH is similar to patterns observed in cancer^9^, and its development and transformation may depend in parts on the altered gene^10–12^, hematopoietic cell-specific rates of mutation, and imposed adaptive pressure on hematopoiesis^13,14^. Cross-sectional studies have demonstrated a higher prevalence of CH mutations, especially at CHIP-defining levels, after exposure to cancer treatment^15^. Moreover, longitudinal characterizations of cancer-free, population-based cohorts as well as patients with solid tumors have elucidated mutation-specific evolution of CH to myeloid malignancies^16–20^. Analyses of CH in patients with solid tumors show distinct mutational patterns and clinical associations under cancer treatment in early and late-stage disease^21–25^; however, evolutionary mechanisms driving differential CH clonal dynamics and its relationship to outcomes are largely unknown.

Extrinsic selection pressures imposed by cancer treatment may differentially induce hematopoietic stress on CH and drive clonal expansion or leukemic transformation, or conversely, lead to clearance of mutated hematopoietic populations after treatment for solid tumors^26–28^. Restricting therapeutic bottlenecks result in deep reductions in hematopoietic allelic populations, establishing an evolutionary setting in which more fit clones are more likely to expand, while less fit or neutral clones may or may not persist due to highly stochastic random drift.

To investigate the impact of cancer treatment on mutation-driven CH, we analyzed 392 serial peripheral blood samples collected from patients with breast cancer before, during, and after their treatment, and determined the impact of therapeutics directed at breast tumor on evolutionary patterns of CH. Our findings, validated in independent cohorts of patients with breast and pan-cancer tumors, quantify the link between CH mutational dynamics and adverse clinical outcomes.

## RESULTS

### Patient population

A total of 171 patients were included in this study based on a confirmed breast cancer diagnosis and the availability of biobanked peripheral blood samples collected both before and after their first treatment for breast cancer (Table 1, Supplementary Figure 1). The median patient age was 58 years (interquartile range [IQR], 50–65 years). All participants were female, with the majority identifying as White (88%) and non-Hispanic (87%). The most common histologic subtype was invasive ductal (67%), followed by invasive lobular (12%) carcinoma. At diagnosis, most patients had early-stage disease and their tumors were hormone receptor-positive (HR+)/HER2- (69%), HER2+ (11%), or triple-negative (13%).

**Table 1:** Clinical characteristics of patients with breast cancer in the discovery cohort

|  | Overall<br>(N=171<sup>1</sup>) | CH<br>(N=94<sup>1</sup>) | No CH<br>(N=77<sup>1</sup>) |
| --- | --- | --- | --- |
| Age | 58 (50, 65) | 62 (55, 67) | 52 (47, 60) |
| Sex |  |  |  |
| Female | 171 (100%) | 94 (100%) | 77 (100%) |
| Race |  |  |  |
| Asian | 3 (1.8%) | 3 (3.2%) | 0 (0%) |
| Black | 10 (5.8%) | 4 (4.3%) | 6 (7.8%) |
| White | 151 (88%) | 85 (90%) | 66 (86%) |
| Other race<sup>2</sup> | 7 (4.1%) | 2 (2.1%) | 5 (6.5%) |
| Ethnicity |  |  |  |
| Hispanic | 23 (13%) | 7 (7.4%) | 16 (21%) |
| Non-Hispanic | 148 (87%) | 87 (93%) | 61 (79%) |
| Smoking status |  |  |  |
| Current | 9 (5.3%) | 7 (7.4%) | 2 (2.6%) |
| Former | 61 (36%) | 36 (38%) | 25 (32%) |
| Never | 101 (59%) | 51 (54%) | 50 (65%) |
| Histology |  |  |  |
| Invasive ductal carcinoma | 114 (67%) | 60 (64%) | 54 (70%) |
| Invasive lobular carcinoma | 21 (12%) | 14 (15%) | 7 (9.1%) |
| Ductal carcinoma in situ | 15 (8.8%) | 10 (11%) | 5 (6.5%) |
| Mixed invasive carcinoma | 11 (6.4%) | 5 (5.3%) | 6 (7.8%) |
| Special subtypes of invasive carcinoma<sup>3</sup> | 10 (5.8%) | 5 (5.3%) | 5 (6.5%) |
| HR/HER2 status<sup>4</sup> |  |  |  |
| HR+/HER2- | 118 (69%) | 61 (65%) | 57 (74%) |
| HR+/HER2+ | 15 (8.8%) | 7 (7.4%) | 8 (10%) |
| HR-/HER2+ | 4 (2.3%) | 4 (4.3%) | 0 (0%) |
| TNBC | 23 (13%) | 14 (15%) | 9 (12%) |
| HR+/HER2 untested | 11 (6.4%) | 8 (8.5%) | 3 (3.9%) |
| Tumor stage at diagnosis, Node, Metastasis stage |  |  |  |
| 0 | 15 (8.8%) | 10 (11%) | 5 (6.5%) |
| 1 | 81 (47%) | 42 (45%) | 39 (51%) |
| 2 | 55 (32%) | 32 (34%) | 23 (30%) |
| 3 | 13 (7.6%) | 7 (7.4%) | 6 (7.8%) |
| 4 | 7 (4.1%) | 3 (3.2%) | 4 (5.2%) |
| Treatment between samples |  |  |  |
| Chemotherapy (± hormone therapy) | 69 (40%) | 33 (35%) | 36 (47%) |
| Radiation therapy (± hormone therapy) | 55 (32%) | 33 (35%) | 22 (29%) |
| Chemotherapy + radiation therapy | 12 (7.0%) | 8 (8.5%) | 4 (5.2%) |
| Hormone only | 35 (20%) | 20 (21%) | 15 (19%) |
| Days between treatment start and first sample | -41 (-66, -14) | -44 (-69, -15) | -41 (-62, -14) |

<sup>1</sup> Median (IQR); n (%).
<sup>2</sup> Other races include patient-reported American Indian or Alaska Native (n=1), Laotian (n=1), or "Other race" (n=5)
<sup>3</sup> Tubular carcinoma, mucinous carcinoma, solid papillary carcinoma with invasion, or metaplastic carcinoma
<sup>4</sup> HR: hormone receptor, + indicates positivity for estrogen and/or progesterone receptor (cutoff $\geq 1\%$ );
TNBC: triple-negative breast cancer

Patients were categorized based on the type of treatment received during the interval between the first and last peripheral blood samples analyzed for CH. Assuming that the influence of hormone therapy on CH is negligible^15^, we classified the patients who received chemotherapy or radiation during the assessed period based on exposure to these modalities. Between the first and last sample, most patients received chemotherapy ± hormone therapy (69/171, 40%) or radiation ± hormone therapy (55/171, 32%). Fewer patients received hormone therapy only (35/171, 20%) or chemotherapy and radiation ± hormone therapy (12/171, 7%) (Table 1).

### CH mutational spectrum at breast cancer diagnosis

To map the mutational landscape of somatic mutations associated with CH, we targeted the full coding regions of 81 genes associated with hematologic malignancies (Supplementary Table 1) using hybrid-capture, error-corrected sequencing at high depth (mean depth of coverage per base=5,679x). Baseline peripheral blood samples were collected 1 day to 7.7 months before the start of treatment. After excluding sequencing errors and likely germline variants, we identified 160 protein-changing CH mutations with variant allele frequencies (VAFs) *>*0.01% in 55% of the cohort (n=94) before treatment start (Figure 1A-B).

**Figure 1:**
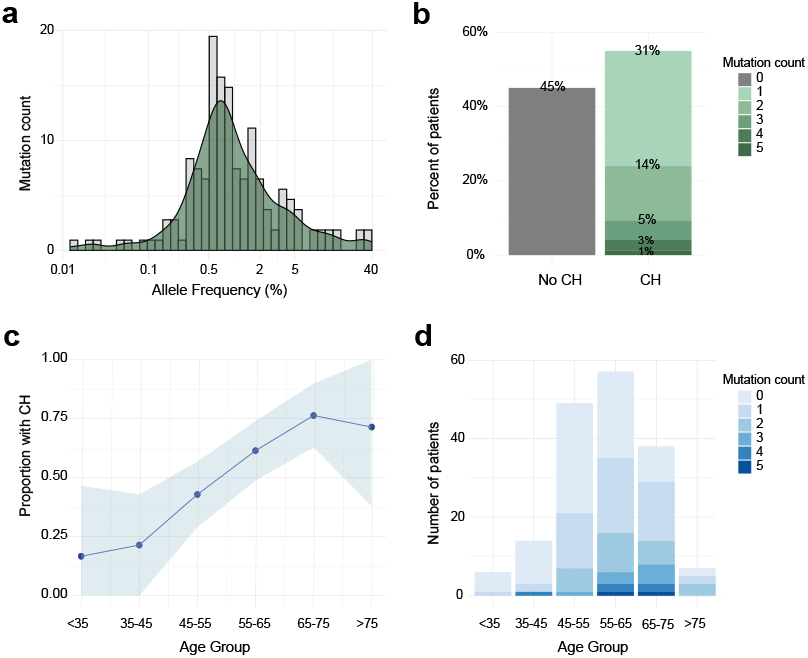
Prevalence and mutational distribution of CH prior to treatment for breast cancer. A) Density distribution of CH mutation variant allele frequencies (VAFs). B) Percent of patients with or without CH, stratified based on the number of detected mutations per individual. C) Proportion of patients with CH across age groups. D) Number of patients with CH across age groups, stratified based on the number of detected mutations per individual.

CH occurs at all ages with a prevalence that begins to rise at age 50–60 years^2,3^. In our cohort, patients with CH at diagnosis were older than patients without CH (median age 62 vs. 52 years, respectively; *P* < 0.001) (Table 1). Accordingly, the proportion of patients with CH increased with age, reaching up to 75% of patients by 65–75 years (Figure 1C). The median number of CH mutations within an individual at breast cancer diagnosis was 1 (range: 0–5) and was associated with age (*P* =0.04) (Figure 1D).

In *DNMT3A, TET2*, and *ASXL1*, the most frequently mutated CH genes in healthy populations^29,30^, we detected missense, loss-of-function nonsense, and frameshift mutations in 65% of the patients with CH (*DNMT3A* n=51/94, 54%; *TET2* n=6/94, 6%; and *ASXL1* n=5/94, 5%) (Figure 2A, Supplementary Table 2). Following *DNMT3A*, the genes with the highest number of protein-changing mutations were *YLPM1* (n=8/94, 8%) and *PPM1D* (n=6/94, 6%). *PPM1D* and *CHEK2* are negative regulators of TP53; accordingly, we identified 4 patients (4%) with mutations in *TP53* affecting its DNA-binding domain and 3 patients (3%) with missense mutations in *CHEK2*.

**Figure 2:**
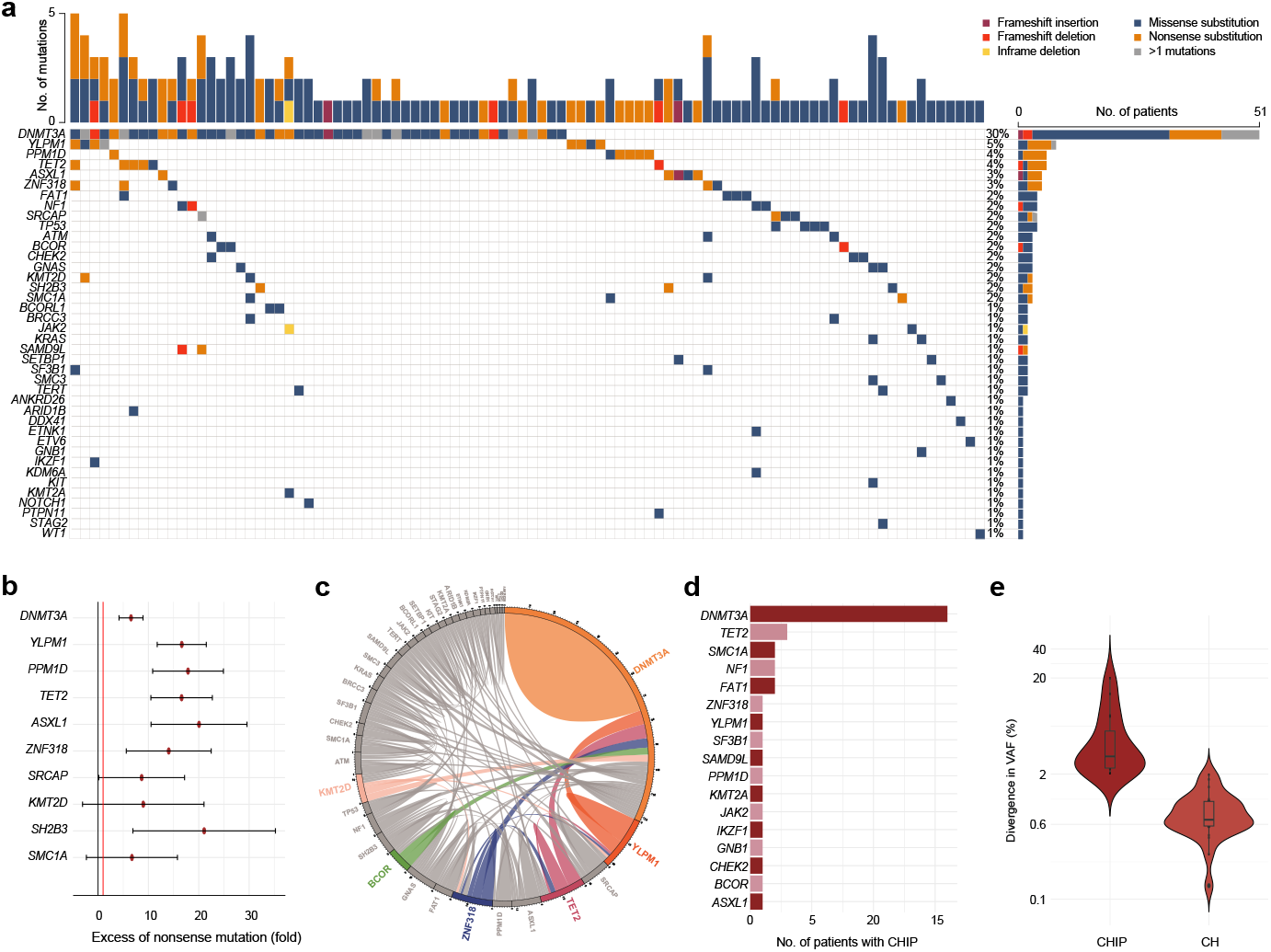
Mutational spectrum and diversity of CH prior to treatment for breast cancer. A) Oncoplot of CH mutations detected across a panel of 81 genes in patients with breast cancer. Columns represent individual patients and colors correspond to the mutation type, including frameshift insertion and deletions, in-frame deletions, and missense and nonsense single nucleotide variants. B) Fold excess of nonsense mutations in each gene relative to the expected distribution from the genes’ nucleotide content; chi-squared statistics shown with 95% confidence intervals for genes with ≥ 3 protein-changing substitutions and ≥ 1 nonsense mutation. C) CH mutation co-occurrence illustrated by a circos plot, linking the most frequently mutated genes and co-mutations. D) Number of patients with CHIP mutations (VAF ≥ 2%) across genes. E) Divergence in CH allelic frequency, defined by mean CH VAF per patient, compared between patients with CHIP mutations (VAF ≥ 2%) versus patients with CH mutations (VAF <2%).

CH mutations arise randomly during hematopoiesis, and some provide a competitive advantage to the harboring hematopoietic stem cells and promote positive clonal expansion^31^. To test whether protein-changing genetic alterations in top mutated genes were under clonal selection, we compared the distribution of all detected CH mutations to the number of missense, nonsense, and synonymous substitutions expected from the genes’ nucleotide content. In 83% (5/6) of genes with at least 5 nonsynonymous mutations, we observed significant 7 to 18-fold excess of nonsense substitutions relative to those expected under neutral selection (FDR<0.001), including in *DNMT3A, YLPM1, PPM1D, TET2*, and *ZNF318* (Figure 2B). *SRCAP* did not show an excess of nonsynonymous mutations. Among the genes with <5 nonsynonymous mutations, we observed a 20-fold excess of nonsense mutations in *ASXL1* (95% CI: 11–30%) and *SH2B3* (95% CI: 7–35%). Although the limited number of missense substitutions in the remaining genes did not provide power to assess their clonal selection (Supplementary Table 3), these results corroborate the extent of population-level, positive selection of loss-of-function mutations in CH^9^.

### CH mutational diversity at breast cancer diagnosis

The clonal allele frequency (*i*.*e*., VAF) and number of CH mutations have been associated with elevated risk of expansion and transformation to malignancy^31^. In our cohort, 56% of patients with CH (n=53/94) harbored only one nonsynonymous mutation before treatment, while 28% had 2 (n=25), 10% had 3 (n=9), 5% had 4 (n=5), and 1% had 5 (n=1) (Figure 1B). The most frequently cooccurring CH mutations involved *DNMT3A*, found co-mutated with *TET2* (n=5), *YLPM1* (n=4), and *ZNF318* (n=3) (Figure 2C). Two *DNMT3A* mutations were detected in eight patients and two *YLPM1* mutations in one additional patient.

The median VAF of all CH mutations was 0.7% (range: 0.01%–38%). When considering only the highest VAF per patient, the median VAF was 1.6% (range: 0.2%–38%). At least one CHIP mutation (*i*.*e*., VAF ≥2%) was present in 33% of patients (n=31/94) before treatment. The most common genes with CHIP mutations before treatment were *DNMT3A* (n=15 patients) and *TET2* (n=3 patients) (Figure 2D). In 4 patients, *DNMT3A*-CHIP co-occurred with at least one other CHIP mutation.

To quantify clonal diversity in somatically mutated populations that arise during CH, we measured divergence in CH VAFs using the standard deviation of mutation VAFs per patient. In patients with *>*1 CH mutation (n=41), the median divergence in CH VAFs was 1% (range: 0.1–20%) and exceeded 2% in 39% of the patients (n=16), all of whom harbored CHIP. In contrast, when no CH met the CHIP threshold, the median divergence in CH VAFs was significantly lower (0.6%, range: 0.1–2.0%, rank-sum *P* =2e-11) (Figure 2E). These results underscore the notion that as early CH clones grow in abundance, a larger number of CH mutations may accumulate and give rise to increased clonal diversity.

### CH mutational rise and fall during breast cancer treatment

To investigate clonal dynamics of CH mutations under breast cancer treatment, we analyzed one to four serial peripheral blood samples (n=230) per patient collected at a median of 5.5 months (range: 0.01–132.3) after treatment start (Figure 3A, Supplementary Figure 2). All alterations detected in posttreatment samples were present in samples collected before treatment, allowing us to track changes in CH VAFs and infer treatment- and gene-specific rates of clonal growth or collapse (Figure 3B).

**Figure 3:**
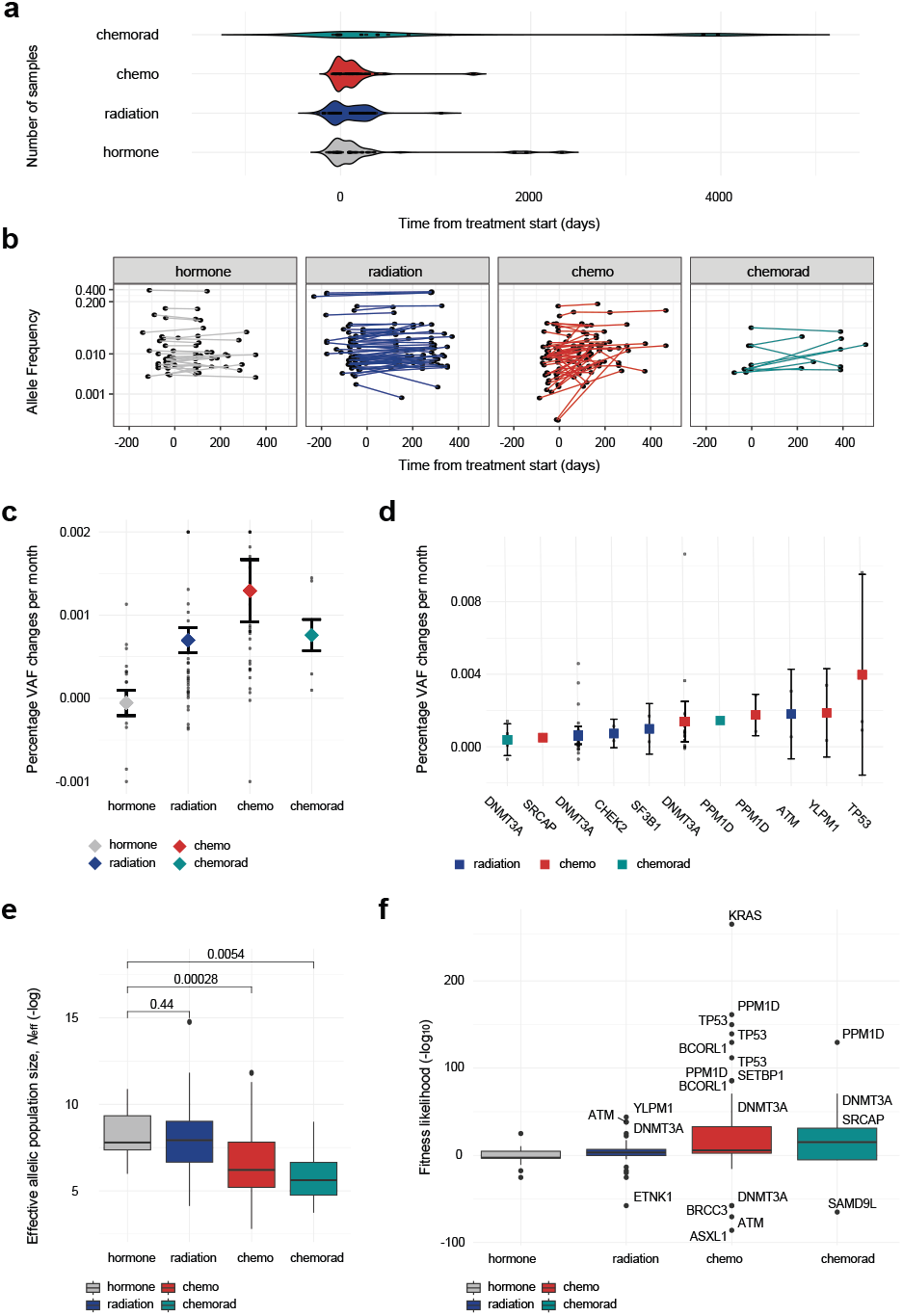
CH mutational dynamic during breast cancer treatment. A) Number and temporal spread of longitudinal samples collected per treatment modality; times are presented relative to treatment start (time=0). B) Change in VAF for all detected CH mutations per treatment modality. C) Percent change in VAF per month for all detected mutations by treatment modality. D) Percent change in VAF per month for gene-specific mutations showing significant allelic increase per treatment modality. E) Effective allelic population size (*N*_eff_) across treatment modalities. F) Fitness likelihood for CH mutations across treatment modalities; genes with mutations showing the highest CH clonal change relative to the wild-type population are indicated.

#### Short-term impact of treatment on CH allele frequency

To evaluate CH temporal dynamics, we first calculated the maximum normalized change in VAF per month for each detected mutation within the first 18 months after treatment start, during which every patient had at least two blood samples collected (Supplementary Table 4). CH VAF increases by treatment were the highest in patients receiving chemotherapy (0.13±0.04% change per month), followed by combination chemotherapy and radiation (0.08±0.02% per month) and radiation only (0.07±0.02% per month) (Figure 3C). There was effectively no measurable VAF change in patients who received only hormonal therapy during the observation period (-0.005±0.02% per month).

The fitness landscape of CH is gene and mutation dependent^9,12,14^. Thus, we calculated gene-specific clonal growth across treatment modalities within the first 18 months after treatment start (Figure 3D, Supplementary Table 4). In patients who were treated with only chemotherapy, CH mutations in *TP53* showed the largest VAF change with an average of 0.40±0.28% increase per month, followed by mutations in *YLPM1* (0.19±0.12% per month), *PPM1D* (0.18±0.06% per month), *DNMT3A* (0.14±0.06% per month), and *SRCAP* (0.05±0.01% per month) (Figure 3D). Mutation-specific increases of *>*0.1% in VAF per month were observed for mutations in *KRAS* (A146V), *BCORL1* (M965I), and *NF1* (M442V) in three patients. Conversely, mutation-specific reductions of *>*0.1% in VAF per month were observed in *FAT1* (E3806K), *ZNF318* (A1519V), *TET2* (A1876V), *ASXL1* (W796X), and *DNMT3A* (E774D, K826R) in six patients and in *BRCC3* (V280D) and *ATM* (M1760V) in one additional patient (Supplementary Table 4).

Treatment with radiation also had a measurable effect on CH growth, where mutations in *ATM* had the highest change with an average of 0.18 ± 0.13% increase in VAF per month (Figure 3D). Mutations in *SF3B1, CHEK2*, and *DNMT3A* showed marginal growth under radiation (0.10±0.07, 0.07±0.04 and 0.06±0.03% increases per month, respectively). Finally, and concordant with our cross-sectional analysis of hormonal therapy’s impact on CH, we did not detect any effective changes in CH VAFs by gene in patients treated with only hormone therapy during the observation period. In fact, *DNMT3A* mutations showed significantly smaller VAF changes in patients treated with hormonal therapy relative to patients under any other treatment modality (rank-sum *P*<0.02) (Supplementary Figure 3A). Sample size was limited to evaluate the impact of combination radiation and chemotherapy per gene; however, we identified two patients in which *DNMT3A* and *PPM1D* mutations had *>*0.1% increase in VAF per month.

All mutations with CHIP-defining VAFs detected before treatment retained CHIP-level VAFs after treatment. Conversely, only 4.4% (n=7/160) of CH mutations expanded to CHIP-defining levels during the same period (*P*<0.005). The CH growth to CHIP occurred in patients receiving chemotherapy (impacting a *TP53* and *JAK2* mutation, and a patient with two *YLPM1* and one *DNMT3A* mutations), radiation (impacting *ASXL1, ATM*, and *DNMT3A* mutations), and combination radiation and chemotherapy (impacting *DNMT3A* and *PPM1D* mutations), but not in patients treated with hormonal therapy (Supplementary Figure 3B).

#### Long-term impact of treatment on CH allelic abundance

To evaluate the extended impact of breast cancer treatment on CH, we analyzed 10 peripheral blood samples from 6 patients collected 22–133 months after treatment start. Across all treatments, CH mutations showed continuous change in VAF from the time of the first sample collection before treatment, with the highest longterm increase in CH VAFs observed in patients treated with cytotoxic chemotherapy. In particular, *DNMT3A* mutations in two patients grew by *>*0.1% in VAF per year over 4 to 8 years of follow-up (Supplementary Table 4).

### CH evolutionary dynamics during breast cancer treatment

When cancer treatment exerts weak selective pressures on CH, it leads to large effective therapeutic bottlenecks that permit the passage of a broader range of clones with comparable fitness. In contrast, strong selective pressures result in more compact therapeutic bottlenecks through which only the most evolutionarily fit clones can pass (Supplementary Figure 1).

#### Effective allelic population during treatment

To quantify therapeutic bottleneck during breast cancer treatment, we assumed that mutations were clonally independent and that VAFs were representative of the prevalence of single clones^9^. We adapted a method for estimating pathogen transmission population size based on allele frequencies^32–34^ and used mutated and wild-type VAFs measured before and after treatment to calculate the lower-bound for the number of alleles that recapitulated the observed mutational dynamics during treatment per patient. This effective allelic population size (*N*_eff_) represents the minimum number of clonally independent alleles needed to explain the observed changes in VAF during treatment in a patient. Therefore, a low *N*_eff_ suggests strong selection and emergence of clonal dominance, while a high *N*_eff_ suggests weaker selection and persistence of greater clonal diversity.

Within the first 18 months of treatment, allelic populations during hormonal therapy were effectively represented by a median of 3,804 alleles (*N*_eff_ range: 397–53,206), comparable to the effective allelic populations estimated under radiation, represented by a median of 2,765 alleles (range: 62–*>*1,000,000). In contrast, and relative to hormonal and radiation, effective allelic populations during chemotherapy were significantly smaller, represented by a median of 496 alleles (range: 16–135,888). With a median of 277 alleles (range: 42–798,800), effective allelic populations under combination radiation and chemotherapy were smaller than under hormonal and radiation, but close to measurements under chemotherapy, pointing to substantially more restrictive therapeutic bottlenecks imposed by cytotoxic treatment of breast cancer (Figure 3E).

#### CH mutational fitness during treatment

The higher a CH mutation’s evolutionary fitness during cancer therapy, the higher the likelihood of its clonal allelic frequency to change as it passes the therapeutic bottleneck. Based on effective allelic population estimates during treatment and using the observed pre- and post-treatment VAFs, we defined a fitness likelihood for each mutation and calculated the probability of its clonal expansion relative to the wild-type alleles. During the first 18 months of treatment, the highest CH mutational fitness was exhibited under chemotherapy. In 27% (22/81) of patients treated with chemotherapy or combination radiation and chemotherapy, the fittest mutated clones per patient were over 10,000 times (*i*.*e*., *>*5 orders of magnitudes) more likely to expand relative to the wild-type population, and harbored mutations in *DNMT3A* (n=7), *TP53* (n=3), *PPM1D* (n=3), *SCRAP* (n=3), *BCORL1* (n=2), *KRAS* (n=1), and *SETBP1* (n=1). Under evolutionary pressures imposed by radiation, *DNMT3A* mutations in three patients and *YLPM1* and *ATM* mutations in single patients had the highest relative fitness and likelihood of growth (Figure 3F). Specifically, the fittest *PPM1D* mutant clones had truncating mutations in exon 6, which have been reported to have a chemoresistance phenotype leading to their expansion with chemotherapy^35^. In addition, 17% of detected *DNMT3A* mutations were positively selected under cytotoxic therapies, all of which were missense or nonsense mutations in the PWWP/chromatin-binding (exons 8 and 9) and DNA methylase (exons 16-19) domains (Supplementary Figures 4).

While we did not observe evidence for recurrent negative selection at the gene level, we measured a significant decrease in VAF of CH mutations under pressures imposed by treatment in 6% (5/81) of patients who received chemotherapy or combination radiation and chemotherapy and 7% (4/55) of patients treated with radiation (Figure 3F, Supplementary Table 5).

#### CH mutational dynamics across treatment modalities and cytotoxic therapy dosage levels

CH has been shown to differentially respond to cancer treatment based on the mechanism of action and dosage. In 38% (36/94) of patients with detectable pre-treatment CH, VAFs showed dynamic changes within the first 18 months of breast cancer treatment, including 9% (3/35) of patients treated with hormonal therapy, 21% (7/55) with radiation, 30% (21/69) with chemotherapy, and 42% (5/12) with combination radiation and chemotherapy. In 3% (3/94) of patients with CH, all of whom received radiation, CH VAFs both increased and decreased. Across all treatment modalities, effective allelic populations were smaller, indicative of stronger selective pressures, in patients with positively or negatively selected CH compared to patients with unchanging CH. Allelic populations contracted most significantly in patients with positively selected CH and treated with chemotherapy or combination radiation and chemotherapy (Figure 4A-B, Supplementary Figure 5A-B), suggesting that therapeutic elimination of wild-type cells may be a major driving factor behind the observed evolutionary fitness of selected CH mutations.

**Figure 4:**
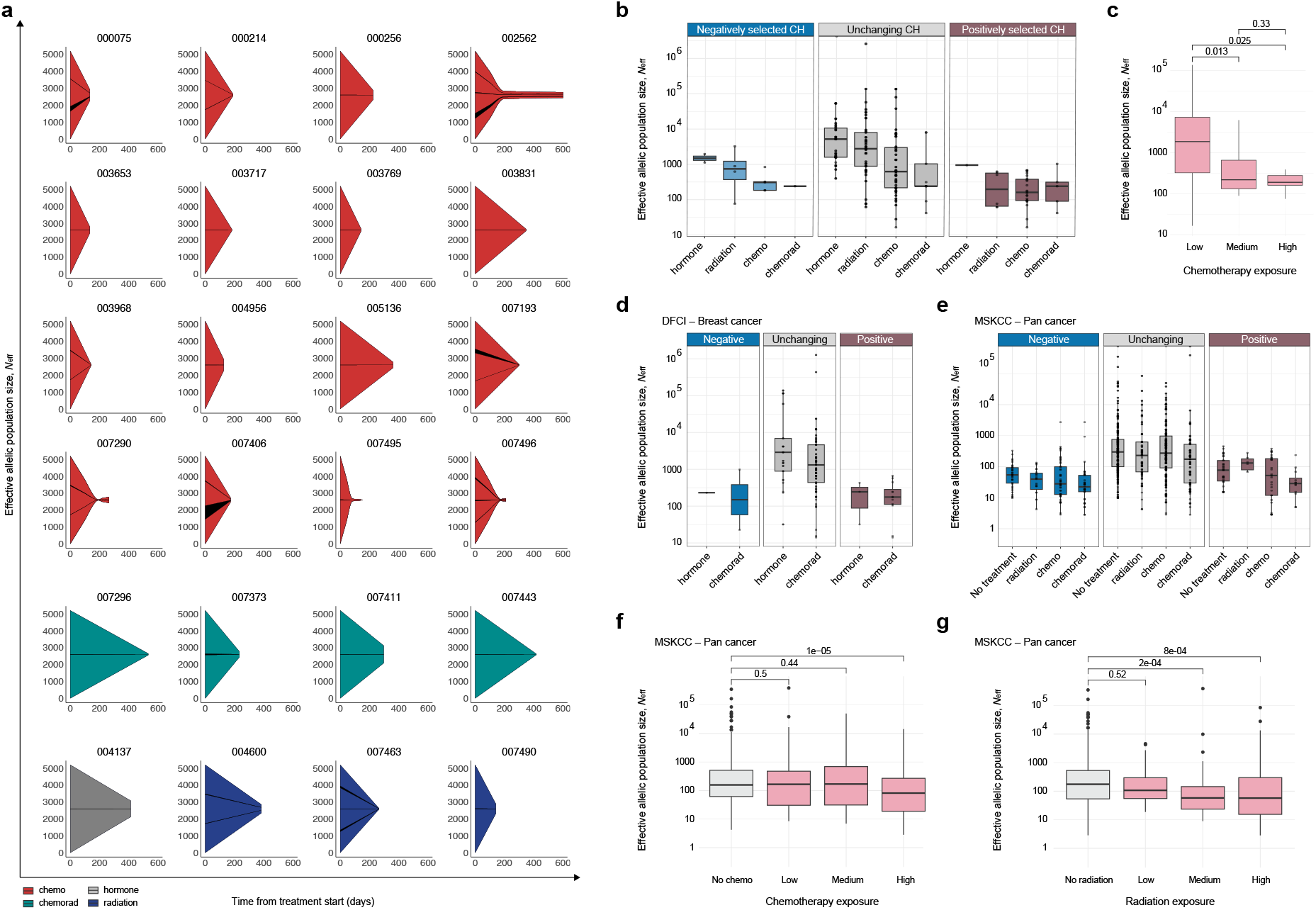
Effective allelic populations associate with treatment modality and dosage. A) Schematics showing effective allelic population size (*N*_eff_) during treatment for patients with positively selected CH, normalized by mean *N*_eff_ in cases treated with hormonal therapy only, across treatment modalities. B) *N*_eff_ stratified based on CH mutational dynamics per patient, including those with positively selected, unchanging, and negatively selected CH. C) *N*_eff_ distributions across cumulative cytotoxic chemotherapy exposure levels. D) *N*_eff_ in patients with breast cancer treated at the Dana Farber Cancer Institute (DFCI) stratified based on CH mutational dynamics per patient, including those with positively selected, unchanging, and negatively selected CH. E) *N*_eff_ in patients with solid tumors treated at Memorial Sloan Kettering Cancer Center (MKSCC) stratified based on CH mutational dynamics per patient, including those with positively selected, unchanging, and negatively selected CH. F) *N*_eff_ distributions across cumulative chemotherapy exposure levels in solid tumors treated at MSKCC. G) *N*_eff_ distributions across cumulative radiotherapy exposure levels in solid tumors treated at MSKCC.

To characterize the impact of therapeutic dosage on CH, we classified patients whose treatment included chemotherapy into three groups with low (40%, n=30/75), medium (41%, n=31/75), or high (19%, n=14/75) cumulative exposure to cytotoxic agents. Likewise, patients treated with radiation were classified into low (40%, n=22/56), medium (30%, n=17/56), or high (30%, n=17/56) cumulative exposure. Patients with CH had similar levels of chemotherapy exposure to patients without CH; thus, we asked whether there was a correlation between the dosage levels and CH mutational dynamics among patients with CH at baseline (n=36). Of those, cumulative chemotherapy exposure was classified as low for 18 patients, medium for 14 patients, high for 4 patients. All 4 patients with high exposure to chemotherapy showed CH dynamics (Supplementary Figure 5C). Moreover, 64% of patients receiving medium and 61% of patients with low levels of cytotoxic chemotherapy exposure had positively or negatively selected CH, pointing to a statistically significant relationship between cytotoxic exposure and CH dynamics (chi-squared *P* =0.003). In addition, we found an association between chemotherapy dosage and effective allelic population size (Figure 4C); patients with high or medium cumulative exposure had significantly smaller effective allelic populations during cytotoxic therapy relative to patients with low-dosage treatment (rank-sum *P* =0.025 and *P* =0.013, respectively). In contrast to systemic chemotherapy, we did not observe a significant relationship between breast-directed radiation exposure levels and allelic population size during treatment. In fact, it is the dose-dependent severity of bottlenecks imposed by systemic cytotoxic therapy that yields shrinking hematopoietic populations which, together with mutation-specific resistance of CH clones to chemotherapeutic drugs^36^, may result in increased CH VAFs during treatment.

### CH mutational dynamics are associated with treatment across all cancers

We tested whether therapy-specific CH mutational dynamics during cancer treatment could be observed in independent groups of patients with breast or other solid tumors. To this end, we analyzed longitudinal CH data from two cohorts of patients who received either or combinations of hormonal therapy, radiation, or chemotherapy including 62 patients with early-stage breast cancer treated at the Dana-Farber Cancer Institute (DFCI)^22^ and 394 patients with solid tumors treated at Memorial Sloan Kettering Cancer Center (MSKCC)^5^.

Corroborating the observations in our discovery cohort, we found stronger selective pressures and smaller effective allelic populations in patients who were under cytotoxic chemo ± radiotherapy compared to those who received hormonal therapy or did not receive any cytotoxic cancer treatment (Figure 4D-E). In patients with early-stage breast cancer (DFCI cohort), allelic populations during radiation and/or chemotherapy were effectively represented by a median of 1,016 alleles (*N*_eff_ range: 14–1,278,372; 47 patients) relative to the effective allelic populations estimated under hormonal therapy, represented by a median of 1,488 alleles (*N*_eff_ range: 32–139,100; 15 patients) (Supplementary Figure 6A), comparable to our discovery cohort. Across all cancers (MSKCC cohort) and relative to the median of 272 alleles (range: 10–352,756) measured for patients who did not receive any treatment, there was a median of 137 alleles (range: 1–8,473) under radiation, 233 alleles (range: 3–49,856) during chemotherapy, and significantly fewer alleles (70, range: 3–391,732) under combination chemotherapy and radiation (rank-sum *P* =2e-04, Supplementary Figure 7A).

As seen with our discovery cohort, effective allelic populations were smaller in patients with positively or negatively selected CH compared to patients with unchanging CH (Figure 4D-E). Moreover, there was a consistent relationship between cytotoxic exposure levels and CH dynamics across all cancers (MKSCC cohort), where positive or negative selection of mutations was significantly associated with higher chemotherapy (chi-squared *P*=0.001) or radiation dosage (chi-squared *P*=1e-04) (Supplementary Figure 7C-D). Accordingly, effective allelic populations were significantly smaller in patients who received the highest cumulative dosage of chemotherapy (rank-sum *P*=1e-05, Figure 4F) or were exposed to high or medium levels of radiation compared to patients who did not receive cytotoxic treatment (rank-sum *P*=2e-04 and *P*=8e-04, respectively, Figure 4G). These results confirm the relationship between cytotoxic treatment dosage and quantified reduction in hematopoietic populations and point to CH clonal dynamics as a biomarker for outcome across cancers^37^.

### CH mutational dynamics are associated with long-term survival and clinical outcomes

CH evolution and growth have been linked with cancer therapy-related adverse sequelae^5,15,18,20^. To evaluate the clinical impact of CH in our cohort, we first compared outcomes for patients with and without CH before treatment and found no significant difference between their overall survival (OS) or progression-free survival (PFS) (Figure 5A). Then we asked whether CH mutational dynamics might reveal associations with clinical outcomes and classified patients with CH into groups with positively selected, negatively selected, and unchanging mutations during breast cancer treatment and found significant differences between overall survival (log-rank *P*=0.0036) (Figure 5B). Most strikingly, survival analysis showed that patients with positively selected CH had an increased risk of death (logrank *P*=4e-04) and a higher risk of progression (log-rank *P*=0.005) relative to patients with negatively selected or unchanging CH (Figure 5C, Supplementary Figure 8). In multivariable Cox regression adjusted for age, stage, and ER, PR, and HER2 status (Supplementary Table 6), positive, compared to negative or unchanging, selection of CH was significantly associated with shorter OS (adjusted Hazard Ratio [HR]=6.6, 95% CI: 1.7–25.9; *P*=0.007) (Figure 5D) and worse PFS (adjusted HR=2.86, 95% CI: 1.01–8.1, *P*=0.049) (Figure 5E). These findings persisted when considering recurrence-free survival and when limited to only patients who received chemotherapy (Supplementary Figure 8C). These results link CH clonal dynamics during treatment to patient mortality and disease outcome, and for the first time, provide quantified evidence for the observed association between mutation-driven CH and OS^31^.

**Figure 5:**
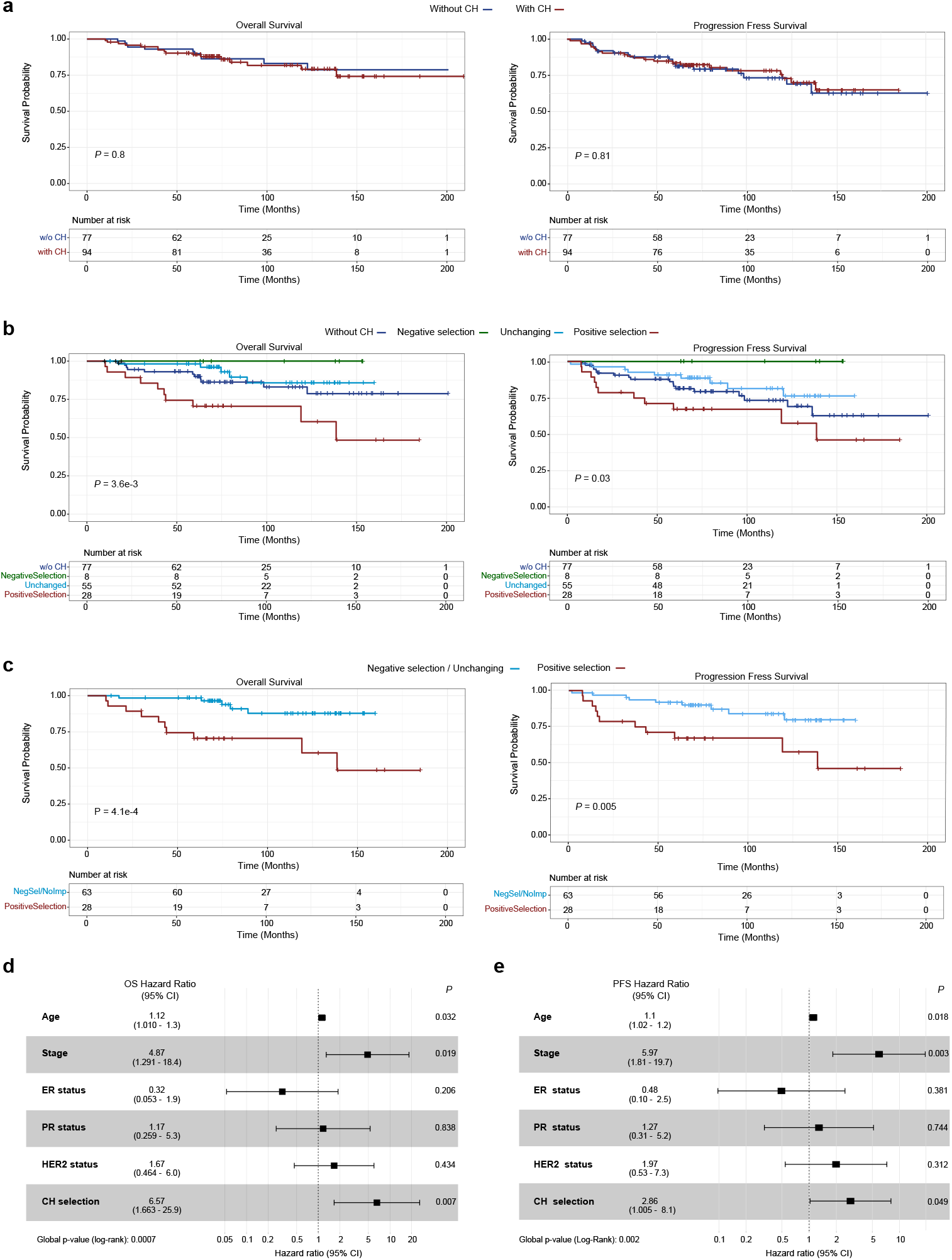
CH mutational dynamics and patient survival. A) Difference in overall survival (OS) and progression-free survival (PFS) between patients with and without CH. B) Differences in OS and PFS between patients with positively selected, negatively selected, unchanging, and no CH. C) Differences in OS and PFS between patients with positively selected CH and those with negatively selected or unchanging CH. D) The impact of positively selected CH on OS using a multivariable Cox regression model including patient age, tumor stage and ER, PR, and HER2 hormonal status. D) The impact of positively selected CH on PFS using a multivariable Cox regression model including patient age, tumor stage and ER, PR, and HER2 hormonal status.

## DISCUSSION

The fitness landscape of CH is shaped by complex, multifactorial processes that include aging, inflammation, germline predisposition^38,39^, and exposure to DNA-damaging agents^31,40^. In this context, cytotoxic treatments in patients with solid tumors have been linked to mechanisms that drive proliferative advantage and, in some cases, transformation to overt hematologic malignancies^15^. In this study, we analyzed serial genomic and clinical data from patients with breast cancer before and after treatment and determined the mutational spectrum, diversity, and distinct, gene-specific evolutionary dynamics of CH under treatment modalities. CH mutations were detected in 55% of patients prior to treatment, an expected prevalence based on the age of the patients and sequencing sensitivity–powered for identifying *>*1 mutant in 1,000 wild-type alleles^10^. Over half of the patients with CH harbored a single mutation, and one-third had mutations with CHIP-defining VAFs. Patients with CHIP mutations had the highest levels of clonal diversity and allelic abundance.

CH mutational spectrum at cancer diagnosis is characterized by an excess of loss-of-function mutations in genes that are under population-level positive selection in healthy individuals^9^. However, CH evolutionary dynamics and degree of associations with adverse outcomes in cancer patients are gene dependent and highly varied^9,12,14^. We assessed mutation-specific fitness during therapeutic-induced hematopoietic stress on CH evolution by analyzing longitudinal samples collected before and after treatment for breast cancer. Notably, all mutations detected after treatment were present before treatment start, demonstrating that no de novo mutations emerged after exposure to treatment for breast cancer. This finding suggests that cancer therapy selectively modulates pre-existing CH rather than initiating development of CH, consistent with reports that CH mutations are not the direct result of DNA-damaging therapeutics and arise due to their context-dependent selective advantage^26^.

While some CH mutations expanded and some contracted during breast cancer treatment, most mutation VAFs did not drastically change (mean change per month=0.04%). Nevertheless, analyzing longitudinal changes in VAFs at high resolution allowed us to dissect the clonal structure of CH and uncover distinct clonal architectures exhibited under different breast cancer treatment modalities. We confirmed previous findings showing minimal impact of hormonal therapy on CH^15^ and found that, in breast cancer, unlike radiation and hormonal therapy, chemotherapy exerts strong selective pressures that significantly reduce effective hematopoietic populations. Consequently, shrinking allelic populations resulted in an effective increase in allelic frequency of persistent CH clones. Mutations in *TP53, PPM1D, SRCAP, DNMT3A*, and *YLPM1* showed the strongest clonal growth under chemotherapy. Mutations in the DNA damage response genes, including *TP53* and *PPM1D*, have been implicated in driving leukemic transformation in therapy-related myeloid neoplasms^5,15,40,41^. Mutations in SRCAP have also been shown to confer selective advantage upon treatment with cytotoxic therapies^42^. Notably, the presence of *YLPM1* as a CH driver that persists and expands with cancer therapy is novel, as most studies do not interrogate this gene; however, both *SRCAP* and *YLPM1* were identified as CH mutations and drivers of myelodysplastic syndromes in large cohorts and showed evidence of positive selection in healthy individuals^9,43^, suggesting their potential role as fitness-enhancing mutations during hematopoietic stress.

Among the most frequent CH drivers, *DNMT3A* mutations selectively expanded under chemotherapy and radiation. Given the link between *DNMT3A* mutations and chemotherapy resistance in hematological malignancies^44^, as well as their association with disease burden and comorbidities in solid tumors^45,46^, these mutations may play a broader role in promoting cancer progression. *DNMT3A* CH frequently co-occurred with *TET2, YLPM1*, and *ZNF318* mutations. While this co-occurrence may suggest clonal cooperation or co-segregation, *TET2* mutations, which are enriched in the tumor microenvironment^7,8^, showed no evidence of positive or negative selection under therapy, indicating they may act as passengers within fitter clones or reflect neutral clonal dynamics. A subset of patients had significant VAF decreases; although no gene-specific trends were noted, this finding shows that there is negative selection of certain CH clones under therapy and underscores the variability in CH clone behavior and the importance of context-specific selection pressures.

Analysis of treatment-specific clonal dynamics showed that chemotherapy, with or without radiation, exerted the strongest selective pressures and resulted in increased CH VAFs. CH dynamics’ link with reduction in effective allelic population size and the significant association with cumulative cytotoxic exposure were independently confirmed in 62 patients with early-stage breast cancer and 394 patients with solid tumors, and reinforce the notion that genotoxic stress promotes relative expansion of clones with higher intrinsic fitness, especially those harboring *TP53* and *PPM1D* mutations. This enrichment aligns with known links between CH and therapy-related neoplasms and may reflect therapy-specific sensitivity or immune-mediated clearance of less fit clones. Of note, the reduced impact of radiation on effective allelic population size and CH selection observed in this study may reflect the limited hematopoietic impact of breast-directed radiation and may not generalize to tumor types where radiation treatment more directly impacts large regions of the hematopoietic stem cell compartment^47^ (*e*.*g*., pelvic radiation for gynecologic cancers^48^). Consistent with this, results of our pan-cancer analysis support the likelihood for dose-dependent radiation-induced CH positive selection in other cancer types.

To explore the impact of CH and clonal evolution on clinical outcomes, we investigated its association with PFS and OS. When classifying patients based on pre-treatment CH status, we did not detect differences in survival, which may be due to our broad classification of CH encompassing low-VAF mutations and/or the overall good long-term survival for patients with breast cancer; however, notable differences arose when considering the evolutionary dynamics of CH mutations with treatment. Specifically, patients with positive selection of their CH clones had an over 6-fold increased risk of mortality compared to patients with negative or neutral selection. While CH dynamics were primarily detected in genes with mutations associated with therapeutic resistance and, therefore, might be confounded by the extent of treatment exposure, we found that patients with negative selection of CH clones had better survival than patients with positive selection and even patients without CH mutations. This suggests that the direction of clonal selection, rather than the mere presence of CH, has prognostic implications. Of note, this effect persisted when the analysis was limited to only patients who received chemotherapy. The mechanism by which positive selection for CH leads to worse outcomes is not clear, although recent data suggest that some CH clones may directly infiltrate tumors and affect tumor growth and treatment response^8,49,50^. Nevertheless, the association between positive selection for CH clones in peripheral blood and outcomes suggests the selected clones have some direct or indirect effects on treatment response.

In conclusion, our study characterizes CH as a dynamic, treatment-sensitive phenomenon with clinically meaningful implications in breast cancer. This is the first longitudinal study to characterize evolutionary dynamics of CH, while prospectively considering independent treatment modalities–chemotherapy, radiation, and hormone therapy–and demonstrating dose-dependent effects. While CH itself may or may not be universally associated with adverse outcomes, the selective expansion of clones with specific mutations may indicate heightened risk for therapy-related complications, progression, or mortality. These results provide strong evidence for clinical relevance of changes in CH allelic abundance during cancer treatment with implications across all solid tumor types.

## METHODS

### Patient population, inclusion criteria, and clinical parameters

Patients for this study were consented to the Moffitt Cancer Center’s Total Cancer Care Protocol, an Institutional Review Board-approved institutional biorepository (MCC#14690; Advarra institutional review board [IRB] Pro00014441)^51^. Use of biobanked patient samples for genetic data generation for this study was approved under a release protocol (MCC#21545, Advarra IRB Pro00058968). Clinical data and samples used for this study were collected between January 1, 1994, and July 31, 2021.

Patients were eligible for inclusion if they had a primary breast cancer diagnosis and serial peripheral blood samples available in our institutional biorepository that could be used for DNA extraction and CH detection, including one sample collected before the start of any cancer treatment and one sample collected after the first therapy given for breast cancer. Specifically, the pre-treatment sample needed to be collected within one year prior to treatment start or a maximum of five days after treatment start for patients treated with chemotherapy or hormone therapy. Patients treated with radiation needed to have a sample collected within a year before radiation start. For patients treated with chemotherapy, the first sequential sample needed to be collected after the chemotherapy stop date, within a year from the start date, and before any radiation treatment. For patients treated with radiation, the first sequential sample needed to be at least 100 days after radiation, within a year from radiation start, and before any chemotherapy treatment. For patients treated with hormone therapy, the first sequential sample had to be collected within a year, but at least 85 days (the average duration of chemotherapy), after hormone therapy start and before any exposure to chemotherapy or radiation. For patients meeting these inclusion criteria, we also included all other serial sampling timepoints they had available in our institutional biorepository, including during treatment and after the post treatment sampling times.

Patients’ cumulative exposure to chemotherapy drugs and radiation during the observation period was calculated following the approach by Bolton et al.^15^, derived from the Late Effects Study Group1^52^. For each patient, cumulative dose per kilogram for each drug received during the observation period was calculated, converted into tertile-based scores, and summed within drug classes. Radiation dose tertiles were calculated using the cumulative radiation dose during the observation period in 2-Gy per fraction (EQD_2_), using an *α/β* of 4 Gy^53^. These class-level scores were then divided into tertiles to categorize overall exposure across the cohort. Of note, cumulative radiation exposure was largely similar across the population (EQD_2_ median, IQR).

### DNA extraction and sequencing

DNA was extracted using the Autopure LS automated DNA extractor (QIAGEN, Hilden, Germany) and quantified using Qubit (Invitrogen, Waltham, MA). Molecular data generation was done in collaboration with Moffitt Molecular Genomics Core. DNA libraries were constructed using a custom designed SureSelect^XTHS2^ kit (Agilent, Santa Clara, CA), which optimally captured whole exons of 84 hematopoietic disorder-related genes, including single nucleotide mutations and small indels. Libraries were sequenced on a NextSeq 2000 sequencer (Illumina, San Diego, CA) per manufacture’s recommendations with a goal of achieving an average depth of coverage *>*5,000x.

### Bioinformatics analysis and somatic variant calling

Sequencing reads were aligned to a corrected human genome (GRCh38) reference^54^ using the BWA-MEM algorithm^55^. Consensus variant calling from reads with the same unique molecular identifiers was done using fgbio v2.2.1^56^. Somatic variant calling was performed using Genome Analysis Toolkit best practices and Mutect2^57^. Additional statistical support for detection of variants in individual samples was obtained using a beta-binomial model for allele-specific sequencing noise (FDR<1e-4)^58^. Variants were filtered and annotated using BCFtools^59^. Insertions and deletions were called by two indel callers, Mutect2 and Lofreq^60^, and were only retained if the VAF was ≥0.02. Variants that occurred in greater than a tenth of the samples were considered sequencing artifacts and were removed. Quality control filters included strand odds ratio <3, quality score *>*6.3, observation of each variant at least once on the forward and reverse reads, and masking of the repetitive regions of the genome as defined by the DUST algorithm^61^ plus a region with low-complexity in *KMT2D* (c. 1659–2544) linked to unreliable calling. Germline variants were removed using publicly available reference populations (i.e., variants observed in non-cancer populations at a prevalence *>*0.005 and with a VAF *>*0.25 were removed^62,63^). To filter for likely functional somatic variants, only mutations or indels located in exonic regions were considered CH mutations. We used germline single nucleotide polymorphism loci heatmaps to confirm correct sample pairing within patients. Chart review confirmed non-tumor cell origin of *TP53* CH mutations and distinction from *TP53* mutations detected by prior sequencing of tumor specimens.

### External validation cohorts

The mutational and clinical data for the DFCI and MSKCC cohorts were obtained through Data Transfer Agreements between the institutions or from their respective publications^5,22^ and were reviewed to match the patient selection and variant calling criteria in the discovery cohort. In particular, we included cases with CH VAFs measured before and after cancer treatment and used exonic variants curated by Vlasschaert et al.^64^ for downstream analyses.

### Clonal selection of CH driver genes

Genes with driver CH mutations were expected to show a higher number of loss-of-function nonsense or hotspot missense mutations compared to expected distribution under neutral selection. Chi-squared statistics were used to compare the observed to expected mutations counts, and lower and upper 95% binomial-approximated confidence intervals were calculated to assess uncertainty for excess of nonsense, missense, and synonymous mutations.

### CH growth rate estimation

Changes in CH allele frequencies and the rate of growth for mutation *j* in patient *i* was defined by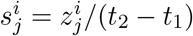, where *t*_1_ and *t*_2_ were collection time points, and 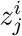 was two-proportion *Z* statistics calculated based on the VAFs (*f*_1_ and *f*_2_) and total sequencing depths (*D*_1_ and *D*_2_) measured in serial samples, following 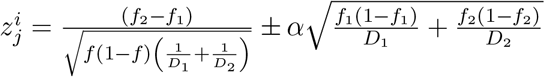 with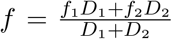. The 95% confidence intervals for growth rate estimates were calculated using confidence intervals for 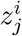 at *α* = 1.96. The mean and standard error in growth rate distributions of mutations were used to calculate growth rates and confidence intervals for specific treatments and genes (per treatment modality).

### Therapeutic bottleneck and evolutionary fitness likelihood calculation

To estimate the effective allelic population size, we assumed that CH mutations were clonally independent and that VAFs were representative of single CH clones. If *n*_*i*_ mutated alleles harbored a specific variant at time point 1, the probability of observing *m*_i_ mutated alleles with the same variant at time point 2 would be described by binomial sampling, 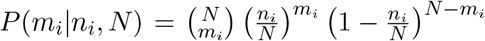, with botlikelihood estimate for *N* tleneck size *N*. For *k* number of CH variants shared between any two time points, the maximum, describing the lower bound on effective bottleneck population size was equal to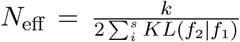, with KL representing the Kullback-Leibler divergence, and *f*_1_ and *f*_2_ as measured VAFs at time points 1 and 2, respectively. The maximum-likelihood-estimated variance of *N*_eff_ was then equal to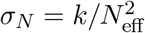.

Using effective allelic population estimates during treatment in each patient, the probability of observing post-treatment VAFs given the CH mutations’ measured pre- and post-treatment VAFs was calculated by 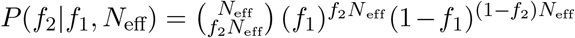. Evolutionary fitness likelihood for mutated and wild-type alleles was then defined by the minus log-likelihood of this probability in base 10.

### Statistical and clinical association analyses

The exact test was used to evaluate cooccurrence of mutations across the gene pairs. Statistically significant differences between groups were calculated for continuous or categorical variables using the exact test, chi-squared, or non-parametric rank-sum test as indicated. The Benjamini-Hochberg false discovery rate (FDR) correction was used for multiple hypotheses testing when indicated.

Clinical associations were determined using a multivariable Cox model regressing clinical values against CH mutational profiles and CH mutational dynamic parameters and states, and by including the following parameters in the model: patient age, breast cancer stage, and ER, PR, and HER2 hormonal status. Statistical analysis and data visualizations were performed using following packages in R: *surminer, survival* (survival analyses), and *ggplot2* (plotting). *Maftools*^65^ was used to generate oncoplot and lollipop figures, and *EvoFreq*^66^ was used to visualize evolutionary models.

## DATA AVAILABILITY

Raw sequencing data of peripheral blood from the discovery cohort (n=171 patients) will be deposited in the dbGaP database prior to publication.

## CODE AVAILABILITY

All code to reproduce and calculate the evolutionary fitness and selection pressure strength are available on GitHub at https://github.com/marabzadeh/EchoCH.

## AUTHOR CONTRIBUTION

NG, HK, and SG conceived and supervised the study. MA performed evolutionary modeling; MA and YT conducted sequencing analyses, variant calling, statistical association analyses, and visualization of the results with help from VM and MT. CCL and JK assisted with identifying the patient cohort. CCL extracted and analyzed clinical data with help from DW and EH. SMa visualized therapeutic bottleneck results with supervision from JW. SMo, HAP, and JEG generated and analyzed the DanaFarber Cancer Institute cohort. All authors drafted the manuscript. All authors read and approved the final manuscript.

## ACKNOWLEDGMENTS

The authors would like to thank Drs. Gregory Riedlinger, Alexandra Jacunski, and Daniel Herranz for the careful reading of the manuscript and for providing constructive feedback.

## SUPPLEMETARY FIGURE LEGENDS

**SUPPLEMETARY FIGURE S1**.

Schematics showing the study design, sampling timeline criteria, and evolutionary models considered for CH during breast cancer treatment.

**SUPPLEMETARY FIGURE S2**.

Swimmer plots showing the sampling timeline relative to breast cancer diagnosis and treatment schedule per patient. A) Patients with overall survival (OS) *≤* 60 months are included. B) Patients with overall survival (OS) 60-120 months are included. C) Patients with overall survival (OS) *>* 120 months are included.

**SUPPLEMETARY FIGURE S3**.

CH mutation-specific changes under treatment for breast cancer. A) Percent change in variant allele frequency (VAF) per month for *DNMT3A* and *YLPM1* mutations by treatment modality. B) Change in VAF for CH mutations that grow to CHIP-defining VAF during treatment. C) Change in VAF for CH mutations in patients with both positive and negative selection.

**SUPPLEMETARY FIGURE S4**.

CH dynamics spectrum. A) The number of variants with positive (Pos) or negative (Neg) selection of CH mutations across treatment modalities and genes. B) Mutational domain spectrum of *DNMT3A* in the 3 groups of negatively selected, positively selected, or unchanging CH.

**SUPPLEMETARY FIGURE S5**.

Schematics showing effective allelic population size (*N*_eff_) during treatment for patients with A) unchanging CH or B) negative selected, normalized by mean *N*_eff_ in cases treated with hormonal therapy only, across treatment modalities. C) Percentage and number of patients across cumulative chemotherapy exposure levels stratified based on CH mutational dynamics.

**SUPPLEMETARY FIGURE S6**.

CH mutational dynamics in the Dana Farber Cancer Institute early-stage breast cancer cohort. A) Effective allelic population size (*N*_eff_) across treatment modalities. B) Percent change in variant allele frequency (VAF) per month for CH mutations by treatment modality. C) REMARK diagram for the DFCI cohort.

**SUPPLEMETARY FIGURE S7**.

CH mutational dynamics in the Memorial Sloan Kettering Cancer Center pan-cancer cohort. A) Effective allelic population size (*N*_eff_) across treatment modalities. A) Percent change in variant allele frequency (VAF) per month for CH mutations by treatment modality. C) Number and percentage of patients across cumulative chemotherapy exposure levels stratified by CH mutational dynamics. D) Number and percentage of patients across cumulative radiotherapy exposure levels stratified by CH mutational dynamics.

**SUPPLEMETARY FIGURE S8**.

Difference in overall survival (OS) and progression free survival (PFS) between patients with positively selected CH and those with either negatively selected or unchanging CH. A) Including only patients with early-stage disease. B) Excluding patients with *TP53* -mutated CH. C) Including patients treated only with chemotherapy during examined period.

## SUPPLEMETARY TABLES

**TABLE S1**.

List of 81 genes included in targeted sequencing

**TABLE S2**.

List of detected CH mutations in the cohort

**TABLE S3**.

Statistics for evaluation of excess of nonsense CH mutations per gene

**TABLE S4**.

Summary of gene level growth rate per treatment

**TABLE S5**.

Statistics for evaluating CH mutation-specific fitness relative to wild-type

**TABLE S6**.

Summary of results for survival analyses using multivariate Cox regression models

## REFERENCES

[1] Alaggio, R. et al. The 5th edition of the world health organization classification of haematolymphoid tumours: lymphoid neoplasms. Leukemia 36, 1720–1748 (2022).

[2] Jaiswal, S. et al. Age-related clonal hematopoiesis associated with adverse outcomes. New England Journal of Medicine 371, 2488–2498 (2014).

[3] Genovese, G. et al. Clonal hematopoiesis and blood-cancer risk inferred from blood DNA sequence. New England Journal of Medicine 371, 2477–2487 (2014).

[4] Ahmad, H., Jahn, N. & Jaiswal, S. Clonal hematopoiesis and its impact on human health. Annual Review of Medicine 74, 249–260 (2023).

[5] Coombs, C. C. et al. Therapy-related clonal hematopoiesis in patients with non-hematologic cancers is common and associated with adverse clinical outcomes. Cell stem cell 21, 374–382. e4 (2017).

[6] Coombs, C. C. et al. Identification of clonal hematopoiesis mutations in solid tumor patients undergoing unpaired next-generation sequencing assays. Clinical Cancer Research 24, 5918–5924 (2018).

[7] Severson, E. A. et al. Detection of clonal hematopoiesis of indeterminate potential in clinical sequencing of solid tumor specimens. Blood, The Journal of the American Society of Hematology 131, 2501–2505 (2018).

[8] Pich, O. et al. Tumor-infiltrating clonal hematopoiesis. New England Journal of Medicine 392, 1594–1608 (2025).

[9] Bernstein, N. et al. Analysis of somatic mutations in whole blood from 200,618 individuals identifies pervasive positive selection and novel drivers of clonal hematopoiesis. Nature Genetics 56, 1147–1155 (2024).

[10] Watson, C. J. et al. The evolutionary dynamics and fitness landscape of clonal hematopoiesis. Science 367, 1449–1454 (2020).

[11] Mitchell, E. et al. Clonal dynamics of haematopoiesis across the human lifespan. Nature 606, 343–350 (2022).

[12] Robertson, N. A. et al. Longitudinal dynamics of clonal hematopoiesis identifies gene-specific fitness effects. Nature medicine 28, 1439–1446 (2022).

[13] Uddin, M. M. et al. Longitudinal profiling of clonal hematopoiesis provides insight into clonal dynamics. Immunity Ageing 19, 23 (2022).

[14] van Zeventer, I. A. et al. Evolutionary landscape of clonal hematopoiesis in 3,359 individuals from the general population. Cancer cell 41, 1017–1031. e4 (2023).

[15] Bolton, K. L. et al. Cancer therapy shapes the fitness landscape of clonal hematopoiesis. Nature genetics 52, 1219–1226 (2020).

[16] Abelson, S. et al. Prediction of acute myeloid leukaemia risk in healthy individuals. Nature 559, 400–404 (2018).

[17] Fabre, M. A. et al. The longitudinal dynamics and natural history of clonal haematopoiesis. Nature 606, 335–342 (2022).

[18] Gillis, N. K. et al. Clonal haemopoiesis and therapy-related myeloid malignancies in elderly patients: a proof-of-concept, case-control study. The lancet oncology 18, 112–121 (2017).

[19] Xie, M. et al. Age-related mutations associated with clonal hematopoietic expansion and malignancies. Nature medicine 20, 1472–1478 (2014).

[20] Takahashi, K. et al. Preleukaemic clonal haemopoiesis and risk of therapy-related myeloid neoplasms: a case-control study. The lancet oncology 18, 100–111 (2017).

[21] Takahashi, K., Nakada, D. & Goodell, M. Distinct landscape and clinical implications of therapy-related clonal hematopoiesis. The Journal of Clinical Investigation 134 (2024).

[22] Morganti, S. et al. Prevalence, dynamics, and prognostic role of clonal hematopoiesis of indeterminate potential in patients with breast cancer. Journal of Clinical Oncology 42, 3666–3679 (2024).

[23] Nead, K. T. et al. Impact of cancer therapy on clonal hematopoiesis mutations and subsequent clinical outcomes. Blood Advances 8, 5215–5224 (2024).

[24] Mayerhofer, C. et al. Clonal hematopoiesis in older patients with breast cancer receiving chemotherapy. JNCI: Journal of the National Cancer Institute 115, 981–988 (2023).

[25] Arends, C. M. et al. Dynamics of clonal hematopoiesis under DNA-damaging treatment in patients with ovarian cancer. Leukemia 38, 1378–1389 (2024).

[26] Bowman, R. L., Busque, L. & Levine, R. L. Clonal hematopoiesis and evolution to hematopoietic malignancies. Cell stem cell 22, 157–170 (2018).

[27] Pich, O. et al. The evolution of hematopoietic cells under cancer therapy. Nature communications 12, 4803 (2021).

[28] Park, S. J. & Bejar, R. Clonal hematopoiesis in cancer. Experimental hematology 83, 105–112 (2020).

[29] Pich, O., Reyes-Salazar, I., Gonzalez-Perez, A. & Lopez-Bigas, N. Discovering the drivers of clonal hematopoiesis. Nature communications 13, 4267 (2022).

[30] Kar, S. P. et al. Genome-wide analyses of 200,453 individuals yield new insights into the causes and consequences of clonal hematopoiesis. Nature Genetics 54, 1155–1166 (2022).

[31] Weeks, L. D. & Ebert, B. L. Causes and consequences of clonal hematopoiesis. Blood 142, 2235–2246 (2023).

[32] Loh, J. W. & Khiabanian, H. Leukemia’s clonal evolution in development, progression, and relapse. Current Stem Cell Reports 5, 73–81 (2019).

[33] Emmett, K. J., Lee, A., Khiabanian, H. & Rabadan, R. High-resolution genomic surveillance of 2014 ebolavirus using shared subclonal variants. PLoS currents 7, ecurrents–outbreaks (2015).

[34] Sobel Leonard, A., Weissman, D. B., Greenbaum, B., Ghedin, E. & Koelle, K. Transmission bottleneck size estimation from pathogen deep-sequencing data, with an application to human influenza A virus. Journal of virology 91, 10.1128/jvi.00171–17 (2017).

[35] Kahn, J. D. et al. PPM1D-truncating mutations confer resistance to chemotherapy and sensitivity to PPM1D inhibition in hematopoietic cells. Blood, The Journal of the American Society of Hematology 132, 1095–1105 (2018).

[36] Hientz, K., Mohr, A., Bhakta-Guha, D. & Efferth, T. The role of p53 in cancer drug resistance and targeted chemotherapy. Oncotarget 8, 8921 (2016).

[37] Cai, R. L.. T. J. J., Xiurong; Bowman. Clonal hematopoiesis in myeloid malignancies and solid tumors. Nat Cancer (2025).

[38] Silver, A. J., Bick, A. G. & Savona, M. R. Germline risk of clonal haematopoiesis. Nature Reviews Genetics 22, 603–617 (2021).

[39] Kessler, M. D. et al. Common and rare variant associations with clonal haematopoiesis phenotypes. Nature 612, 301–309 (2022).

[40] Florez, M. A. et al. Clonal hematopoiesis: Mutation-specific adaptation to environmental change. Cell stem cell 29, 882–904 (2022).

[41] Hsu, J. I. et al. PPM1D mutations drive clonal hematopoiesis in response to cytotoxic chemotherapy. Cell stem cell 23, 700–713. e6 (2018).

[42] Chen, C.-W. et al. SRCAP mutations drive clonal hematopoiesis through epigenetic and dna repair dysregulation. Cell Stem Cell 30, 1503–1519. e8 (2023).

[43] Beauchamp, E. M. et al. ZBTB33 is mutated in clonal hematopoiesis and myelodysplastic syndromes and impacts rna splicing. Blood cancer discovery 2, 500–517 (2021).

[44] Guryanova, O. A. et al. DNMT3A mutations promote anthracycline resistance in acute myeloid leukemia via impaired nucleosome remodeling. Nature medicine 22, 1488–1495 (2016).

[45] Feng, Y. et al. Hematopoietic-specific heterozygous loss of dnmt3a exacerbates colitis-associated colon cancer. Journal of Experimental Medicine 220, e20230011 (2023).

[46] Yan, B. et al. Clonal hematopoiesis driven by dnmt3a mutations promotes metabolic disease development. bioRxiv 2025.05. 02.651715 (2025).

[47] Goglia, A. G. et al. Impact of radiotherapy site, modality, and dose on subsequent clonal hematopoiesis. Blood Advances (2025).

[48] Corbeau, A. et al. Correlations between bone marrow radiation dose and hematologic toxicity in locally advanced cervical cancer patients receiving chemoradiation with cisplatin: a systematic review. Radiotherapy and Oncology 164, 128–137 (2021).

[49] Buttigieg, M. M., Vlasschaert, C., Bick, A. G., Vanner, R. J. & Rauh, M. J. Inflammatory reprogramming of the solid tumor microenvironment by infiltrating clonal hematopoiesis is associated with adverse outcomes. Cell Reports Medicine 6 (2025).

[50] Vanner, R. J. et al. Hematopoietic tet2 inactivation enhances the response to checkpoint blockade immunotherapy. bioRxiv 2024.09. 09.612140 (2024).

[51] Fenstermacher, D. A., Wenham, R. M., Rollison, D. E. & Dalton, W. S. Implementing personalized medicine in a cancer center. The Cancer Journal 17, 528–536 (2011).

[52] Tucker, M. A. et al. Leukemia after therapy with alkylating agents for childhood cancer. Journal of the National Cancer Institute 78, 459–464 (1987).

[53] Whelan, T. J.Kim, D.-H. & Sussman, J. Clinical experience using hypofractionated radiation schedules in breast cancer. In Seminars in radiation oncology, vol. 18, 257–264 (Elsevier).

[54] Miller, C. A. et al. Failure to detect mutations in U2AF1 due to changes in the GRCh38 reference sequence. The Journal of Molecular Diagnostics 24, 219–223 (2022).

[55] Li, H. Aligning sequence reads, clone sequences and assembly contigs with BWA-MEM (2013).

[56] Fennel, T., Homer, N. & Genomics, F. Fgbio: tools for working with genomic and high throughput sequencing data. URL https://github.com/fulcrumgenomics/fgbio. (accessed July 2025).

[57] McKenna, A. et al. The genome analysis toolkit: a mapreduce framework for analyzing next-generation dna sequencing data. Genome research 20, 1297–1303 (2010).

[58] Rabadan, R. et al. On statistical modeling of sequencing noise in high depth data to assess tumor evolution. Journal of statistical physics 172, 143–155 (2018).

[59] Li, H. A statistical framework for SNP calling, mutation discovery, association mapping and population genetical parameter estimation from sequencing data. Bioinformatics 27, 2987–2993 (2011).

[60] Wilm, A. et al. LoFreq: a sequence-quality aware, ultra-sensitive variant caller for uncovering cell-population heterogeneity from high-throughput sequencing datasets. Nucleic acids research 40, 11189–11201 (2012).

[61] Morgulis, A., Gertz, E. M., Schäffer, A. A. & Agarwala, R. A fast and symmetric DUST implementation to mask low-complexity DNA sequences. Journal of computational biology 13, 1028–1040 (2006).

[62] Karczewski, K. J. et al. The mutational constraint spectrum quantified from variation in 141,456 humans. Nature 581, 434–443 (2020).

[63] Mailman, M. D. et al. The NCBI dbGaP database of genotypes and phenotypes. Nature genetics 39, 1181–1186 (2007).

[64] Vlasschaert, C. et al. A practical approach to curate clonal hematopoiesis of indeterminate potential in human genetic data sets. Blood, The Journal of the American Society of Hematology 141, 2214–2223 (2023).

[65] Mayakonda, A., Lin, D.-C., Assenov, Y., Plass, C. & Koeffler, H. P. Maftools: efficient and comprehensive analysis of somatic variants in cancer. Genome research 28, 1747–1756 (2018).

[66] Gatenbee, C. D., Schenck, R. O., Bravo, R. R. & Anderson, A. R. EvoFreq: visualization of the evolutionary frequencies of sequence and model data. BMC bioinformatics 20, 710 (2019).

