## Supplemental Figures for "Clonal Hematopoiesis Dynamics and Evolutionary Fitness During Cancer Treatment Impact Clinical Outcomes"

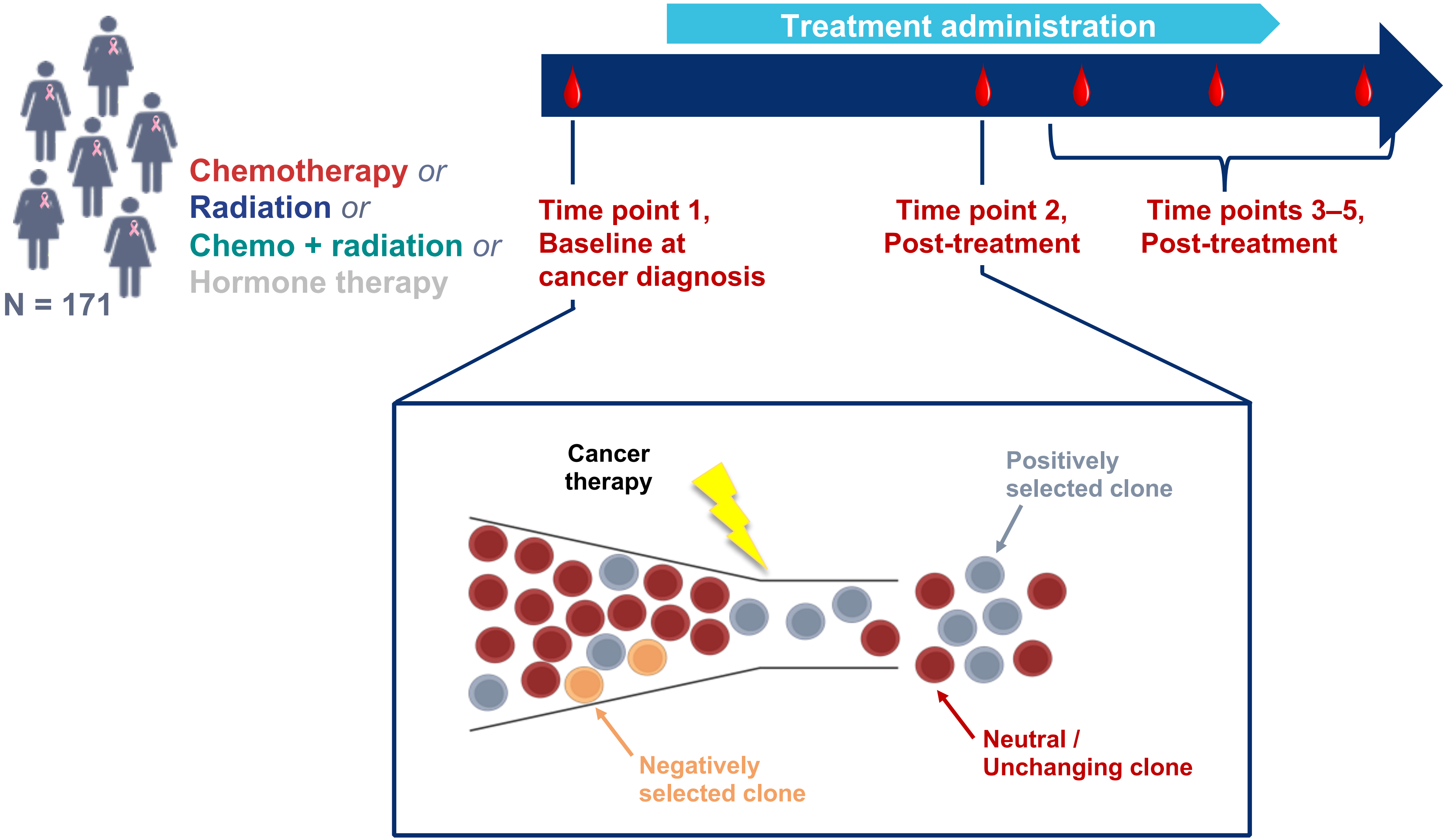


**FIGURE S1.** Schematics showing the study design, sampling timeline criteria, and evolutionary models considered for CH during breast cancer treatment.

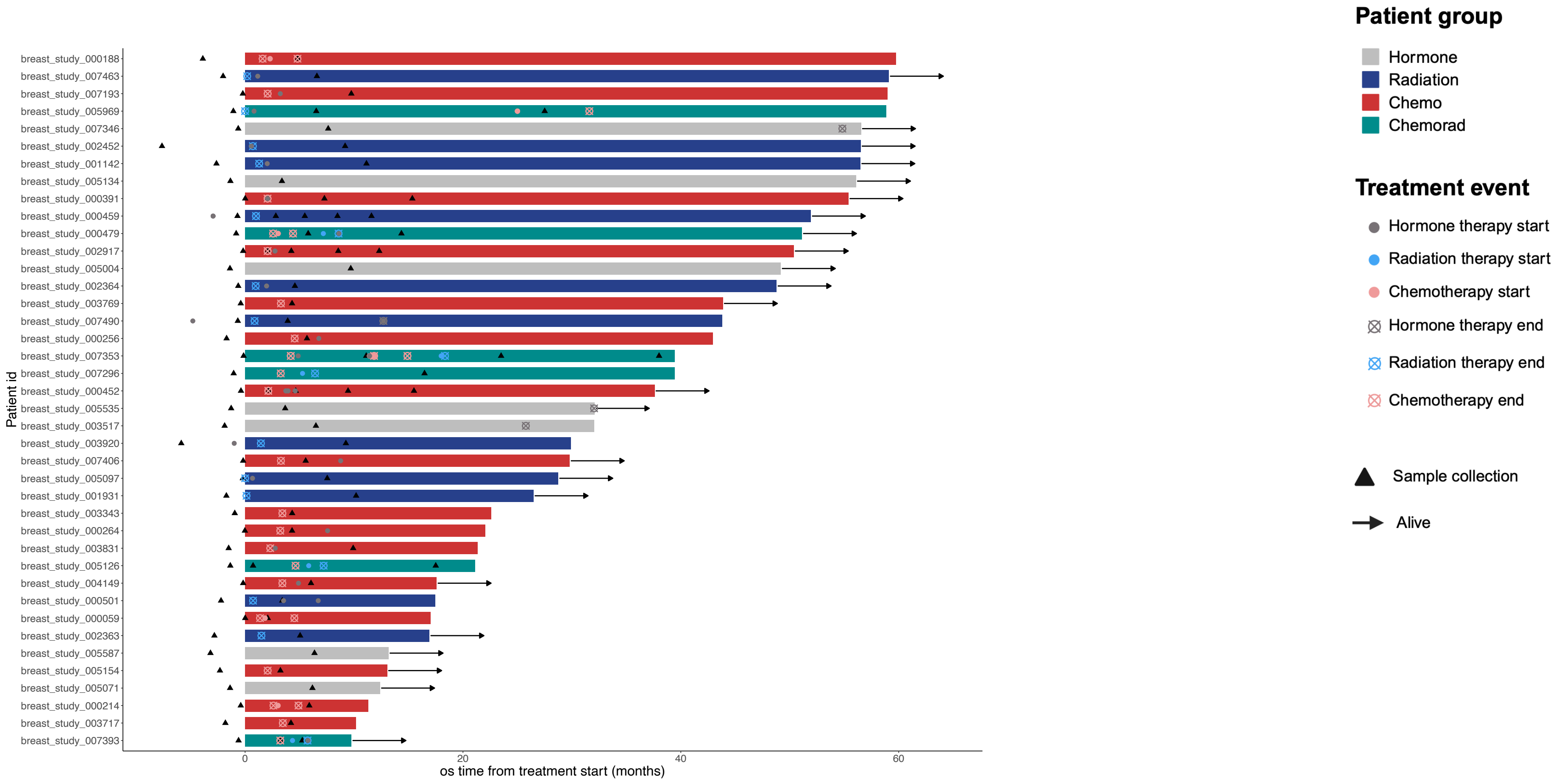


**FIGURE S2A.** Swimmer plots showing the sampling timeline relative to breast cancer diagnosis and treatment schedule per patient. Patients with overall survival (OS) ≤ 60 months are included.

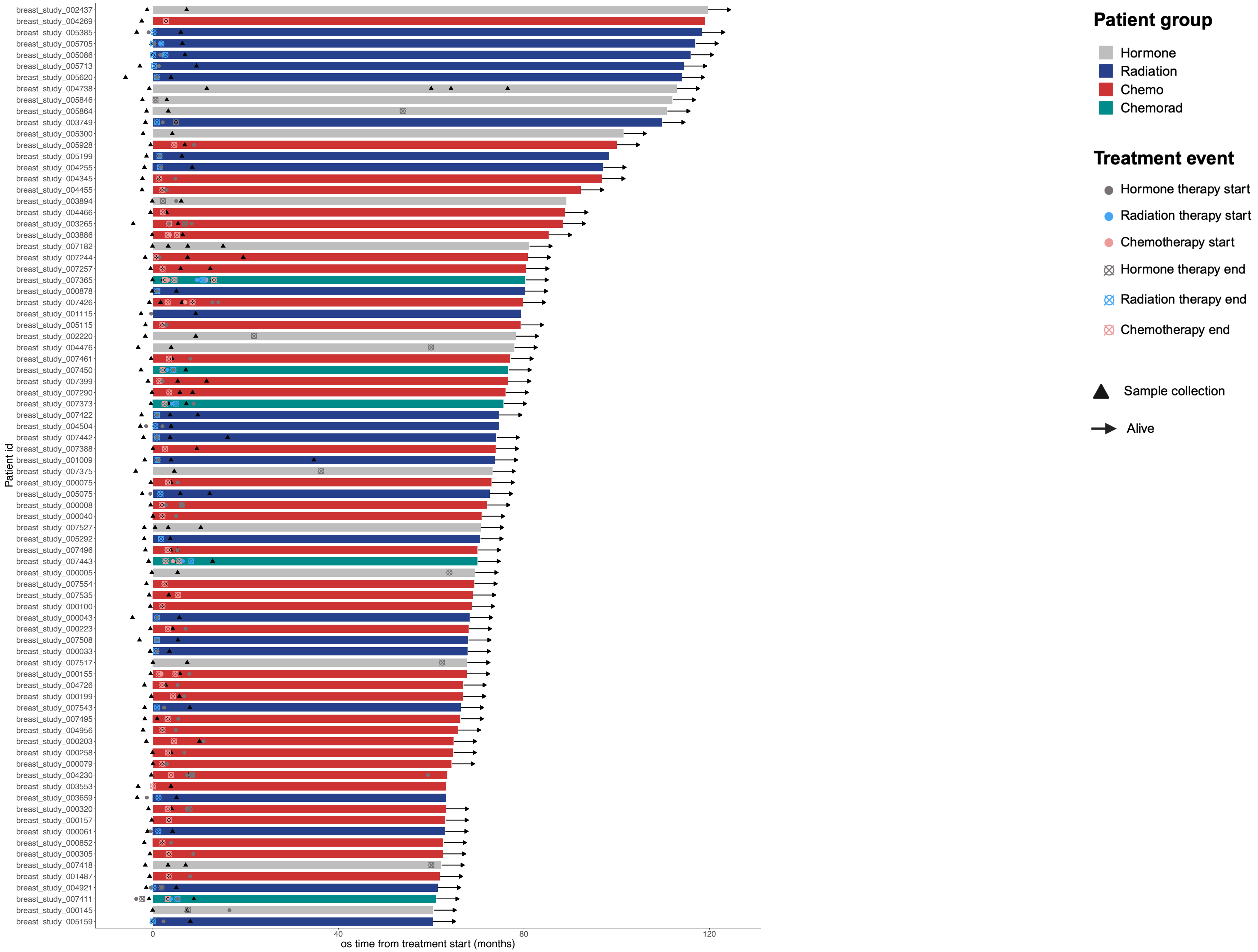


**FIGURE S2B.** Swimmer plots showing the sampling timeline relative to breast cancer diagnosis and treatment schedule per patient. Patients with overall survival (OS) 60-120 months are included.

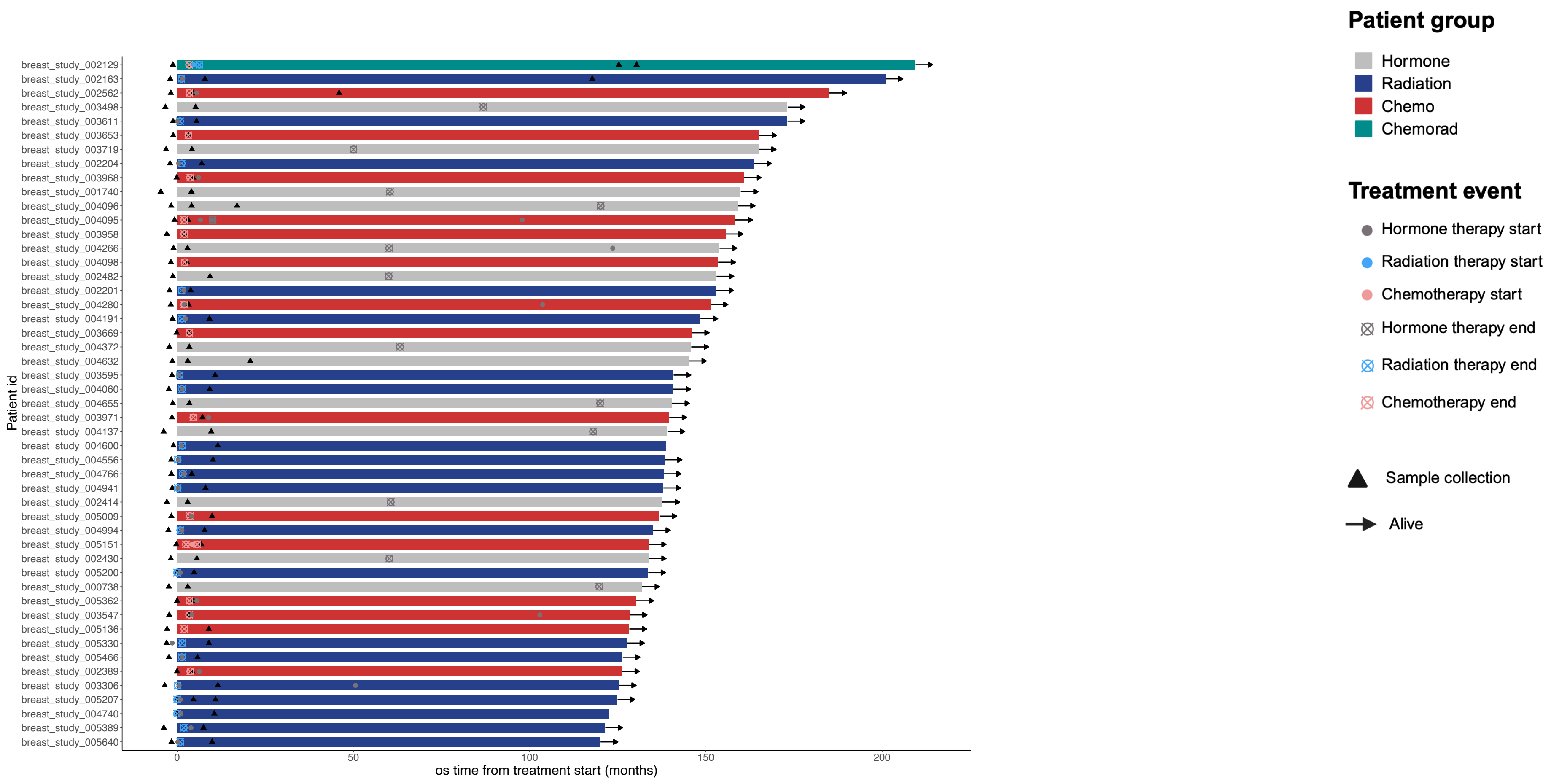


**FIGURE S2C.** Swimmer plots showing the sampling timeline relative to breast cancer diagnosis and treatment schedule per patient. Patients with overall survival (OS) > 120 months are included.

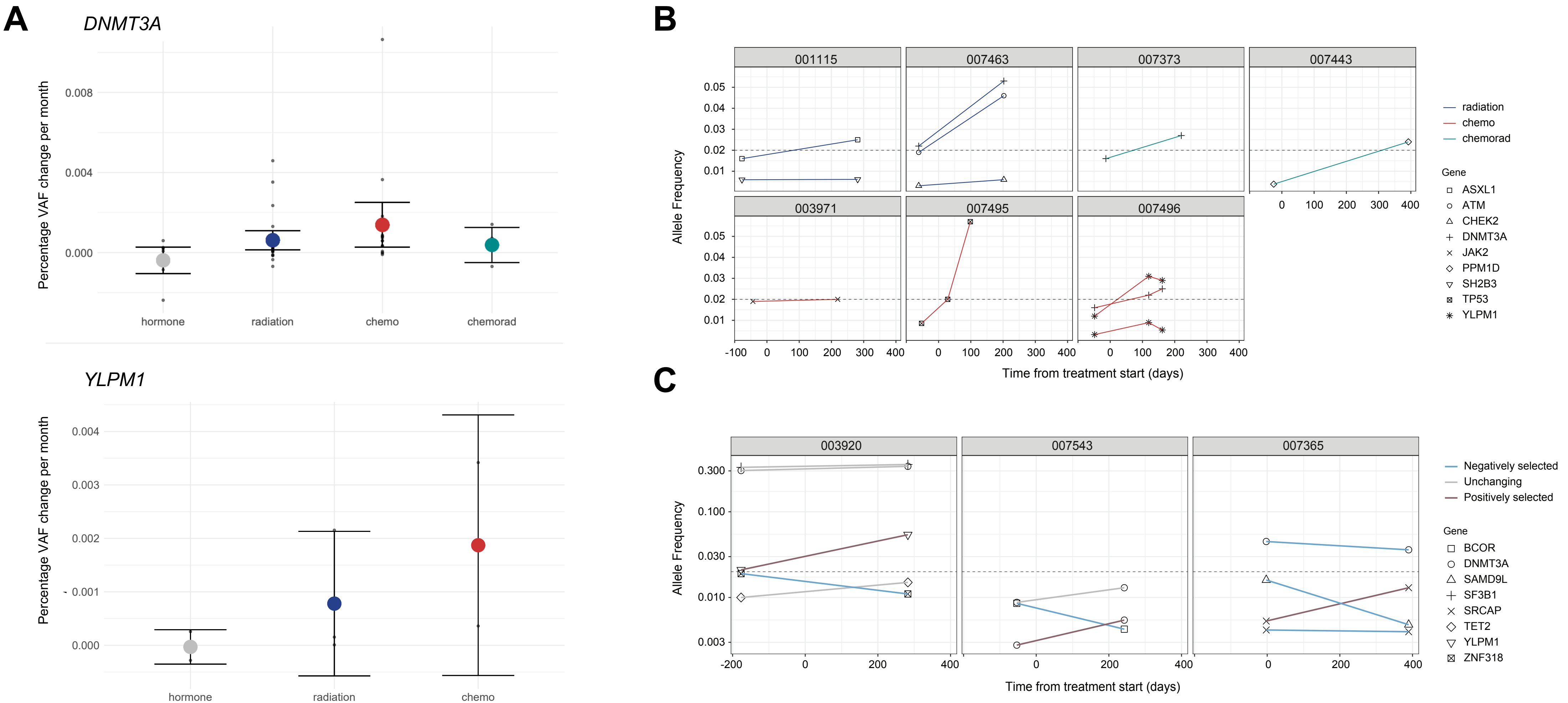


**FIGURE S3.** CH mutation-specific changes under treatment for breast cancer. A) Percent change in variant allele frequency (VAF) per month for DNMT3A and YLPM1 mutations by treatment modality. B) Change in VAF for CH mutations that grow to CHIP-defining VAF during treatment. C) Change in VAF for CH mutations in patients with both positive and negative selection.

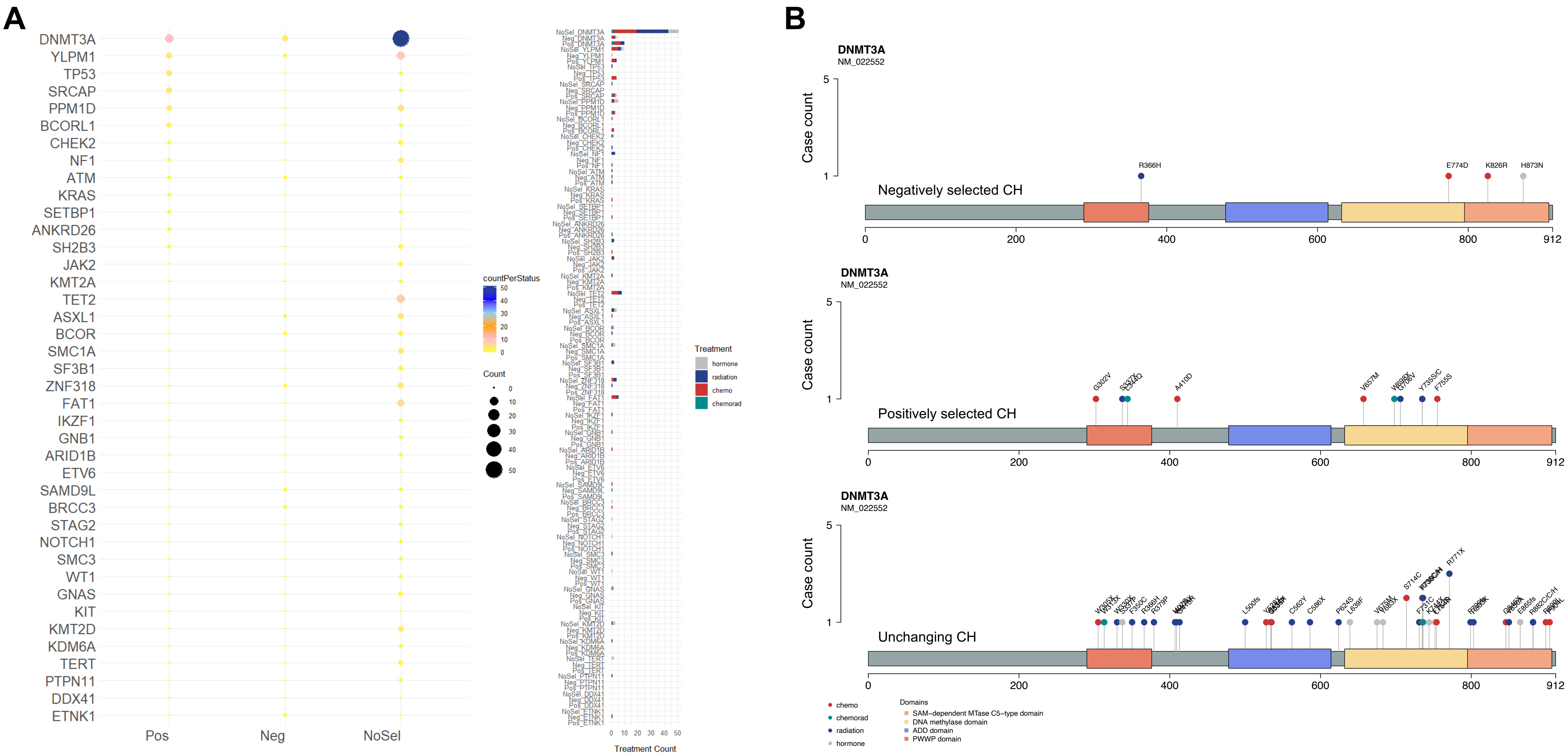


**FIGURE S4.** CH dynamics spectrum. A) The number of patients with positive (Pos), negative (Neg), or no selection (NoSel) of CH mutations across treatment modalities and genes. B) Mutational domain spectrum of DNMT3A in the 3 groups of negatively selected, positively selected, or unchanging CH.

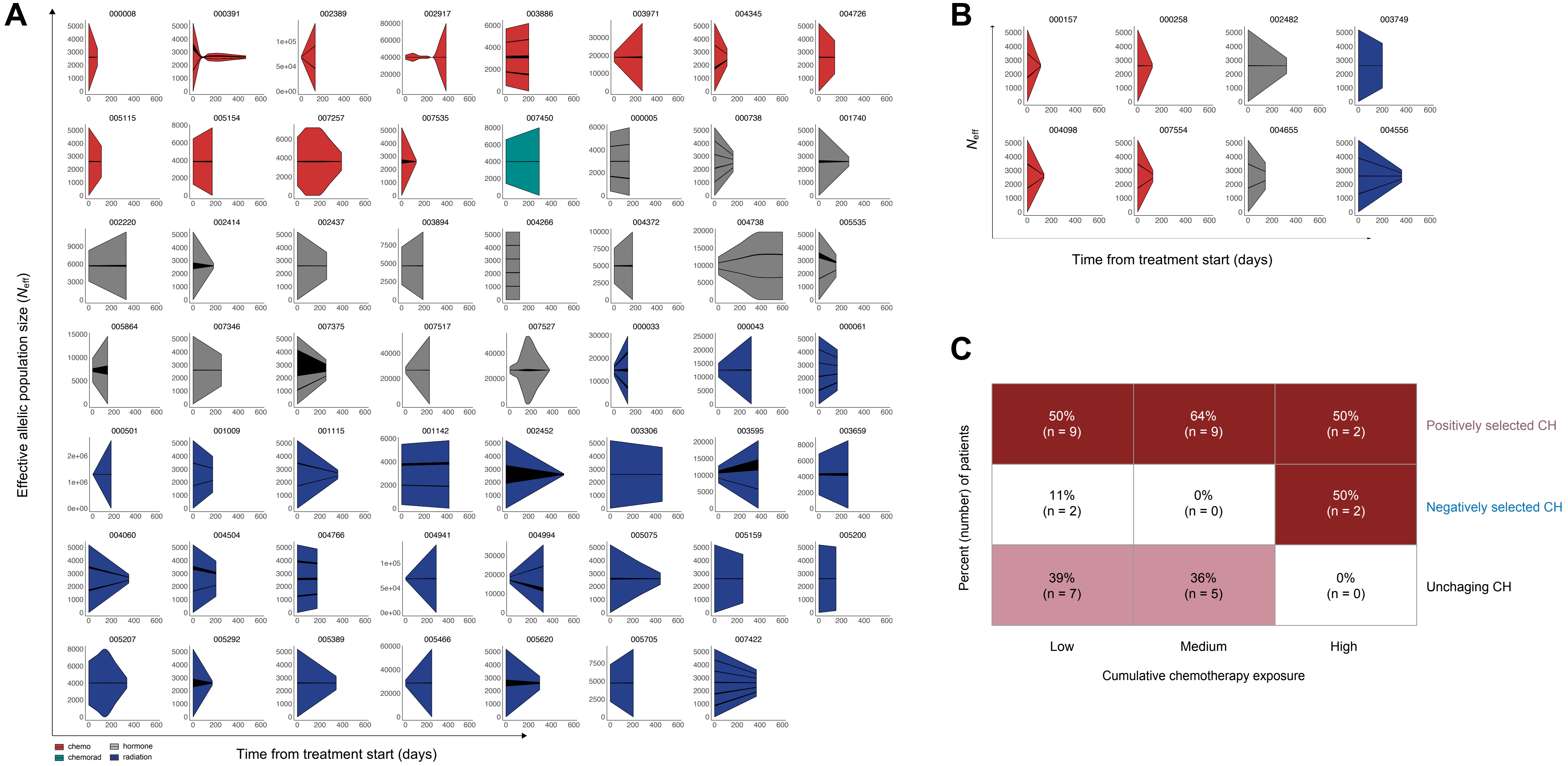


**FIGURE S5.** Schematics showing effective allelic population size (Neff) during treatment for patients with A) unchanging CH or B) negative selected CH, normalized by mean $N_{eff}$ in cases treated with hormonal therapy only, across treatment modalities. C) Percentage and number of patients across cumulative chemotherapy exposure levels stratified based on CH mutational dynamics.

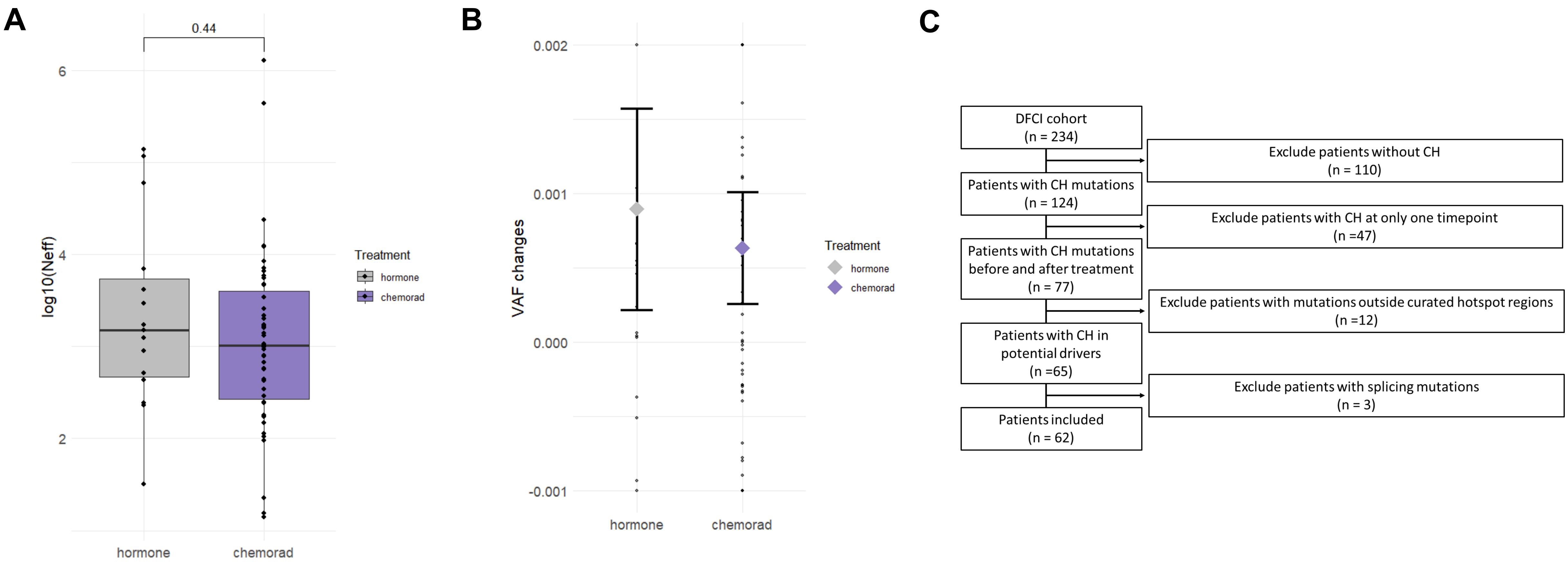


**FIGURE S6.** CH mutational dynamics in the Dana Farber Cancer Institute early-stage breast cancer cohort. A) Effective allelic population size (Neff) across treatment modalities. B) Percent change in variant allele frequency (VAF) per month for CH mutations by treatment modality. C) REMARK diagram for the DFCI cohort.

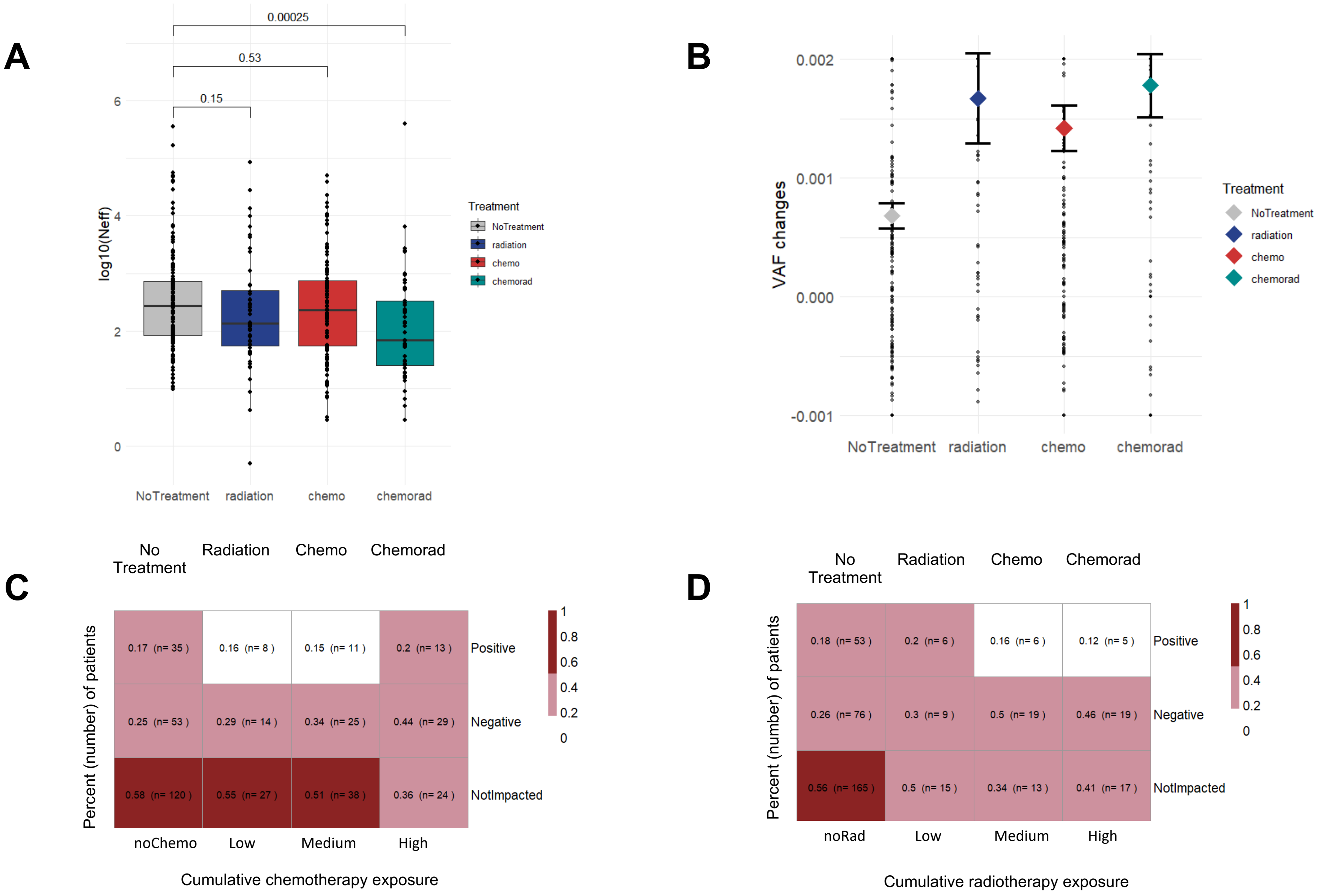


**FIGURE S7.** CH mutational dynamics in the Memorial Sloan Kettering Cancer Center pan-cancer cohort. A) Effective allelic population size (Neff) across treatment modalities. A) Percent change in variant allele frequency (VAF) per month for CH mutations by treatment modality. C) Number and percentage of patients across cumulative chemotherapy exposure levels stratified by CH mutational dynamics. D) Number and percentage of patients across cumulative radiotherapy exposure levels stratified by CH mutational dynamics.

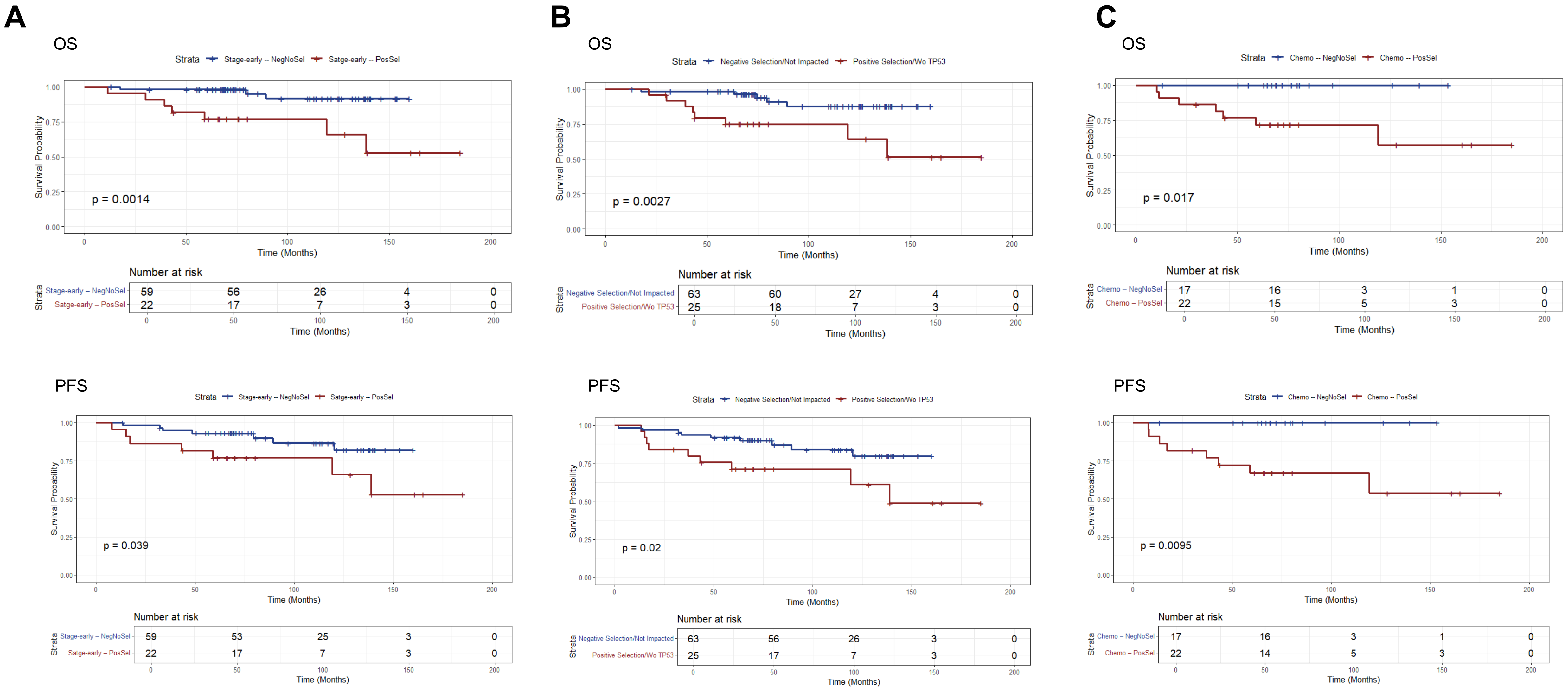


**FIGURE S8.** Difference in overall survival (OS) and progression free survival (PFS) between patients with positively selected CH and those with either negatively selected or unchanging CH. A) Including only patients with early-stage disease. B) Excluding patients with TP53-mutated CH. C) Including patients treated only with chemotherapy during examined period.
